# Interactions between Insomnia and Obstructive Sleep Apnea

**DOI:** 10.64898/2026.07.12.26357841

**Authors:** Yanyuan Dai, Yun Li, Elisabeth Heremans, Ulysse Gimenez, Umaer Hanif, Emmanuel Mignot

## Abstract

**Study Objectives:** Co-morbid insomnia and sleep apnea (COMISA) is challenging clinically and difficult to treat. Our goal was to assess how much COMISA is the mere addition of two phenotypes or display features indicative of genuine statistical interactions.

**Methods:** A total of 152,487 patients from 240 sleep centers across 30 US states were included. Insomnia was defined as difficulty initiating/maintaining sleep with daytime fatigue/sleepiness occurring “often”/“always”. OSA was defined as having an Apnea Hypopnea Index (AHI) more than 15 events/h. Modified Poisson regression was conducted to evaluate multiplicative interactions between insomnia and OSA on common comorbidities and sleep symptoms. Additive interactions were also examined. Linear regression models were used to evaluate additive interactions for PSG parameters. The false discovery rate was controlled using the Benjamini–Hochberg procedure.

**Results:** After adjustment for confounders, insomnia and OSA demonstrated positive interactions for depression, chronic muscular pain, headache, subjective excessive daytime sleepiness (EDS), naps, and pre-sleep anxious and muscular tension (adjusted p < 0.05). Furthermore, insomnia and OSA demonstrated positive interactions for parameters related to respiratory disturbance, including AHI, oxygen desaturation index (ODI), respiratory disturbance index (RDI), total arousal index (AI) and respiratory AI, and negative interactions for minimum oxygen saturation and percentage of rapid eye movement stage (REM%) (adjusted p < 0.05). Furthermore, the adverse effects of insomnia and OSA on AHI, ODI, RDI and REM% were substantially amplified in males.

**Conclusions:** Our findings demonstrate that insomnia and OSA do not merely coexist but genuinely interact synergistically to amplify selected adverse clinical outcomes.

**Statement of Significance:** Co-morbid insomnia and sleep apnea (COMISA) has a particular symptomatic presentation and is difficult to treat. Researchers have suggested there is a bidirectional causal relationship between the two disorders based on shared pathophysiology. Pursuing on this hypothesis, we explore statistical interactions between insomnia and OSA on common comorbidities, sleep symptoms, and PSG parameters in a large multi-center clinical sample. We provide for the first time evidence that insomnia and OSA truly interact to exacerbate a broad spectrum of clinical outcomes, including depression, chronic muscular pain, headache, subjective EDS, pre-sleep cognitive-emotional and somatic hyperarousal, physiological respiratory disturbances, and sleep disruption. These findings indicate that insomnia and OSA do not merely coexist but potentiate each other’s adverse health effects.

## Introduction

Insomnia and obstructive sleep apnea (OSA) are the two most prevalent sleep disorders worldwide. Not surprisingly, they frequently co-exist, with 30-40% of patients with insomnia having co-morbid OSA, and 30-50% of patients with OSA having co-morbid insomnia[1]. According to the latest meta-analysis, which included 16 studies, the prevalence of co-morbid insomnia and sleep apnea (COMISA) is about 30%[2]. COMISA prevalence estimates vary according to the definitions of insomnia and OSA as well as the population under investigation[3]. Using an apnea hypopnea index (AHI) threshold more than 15 events/h to define moderate-to-severe OSA, previous studies have reported a COMISA prevalence of approximately 13.9% among insomnia sufferers[4], compared with only about 2.6% in the general population[5]. Patients with COMISA present unique and complex diagnostic and treatment needs compared to patients with insomnia or OSA alone[6, 7].

Historically, insomnia and OSA have been regarded as two separate sleep disorders, each with its distinct manifestations, symptoms, pathogenesis, diagnostic and treatment requirements. Insomnia is characterized by frequent occurrences of difficulty falling asleep, maintaining sleep, and/or early morning awakenings, accompanied by a significant daytime impairment. It is more frequent in women, shows an association with depression and is thought to result from maladaptive cognitive learning and/or physiological hyperarousal, with resulting activation of the sympathetic nervous system[8–11]. OSA is more frequent in men, post-menopausal women and associated with obesity[12, 13]. It is characterized by repeated episodes of reduced airflow and/or cessation of breathing during sleep, leading to intermittent hypoxia and sleep fragmentation, and is commonly accompanied by daytime dysfunction, notably daytime sleepiness[14, 15]. Multiple studies have shown associations with cardiovascular disease independent of Body Mass Index (BMI)[16–18].

COMISA was first described by Guilleminault and colleagues in 1973[19], however, it has only gained progressive recognition over the past two decades. Recently, COMISA has emerged as a clinically important phenotype characterized by increased clinical burden across multiple health domains[20]. Accumulating evidence suggests that COMISA is associated with a substantially increased cardiometabolic and psychiatric comorbidities[20–22]. Compared with either disorder alone, COMISA has been linked to a higher risk of hypertension, cardiovascular disease, metabolic syndrome[3, 23, 24]. In addition, population-based and clinical studies have shown that patients with COMISA exhibit significantly higher levels of depressive symptoms, anxiety, and psychological distress than those with either disorder alone[25–28]. More importantly, emerging longitudinal evidence further suggests that COMISA was associated with an elevated risk of all-cause mortality[29, 30].

The pathophysiology of COMISA involves complex and interconnected mechanisms. A meta-analysis examined the pathophysiological correlations of COMISA suggested that differences are particularly evident in the indicators AHI, minimum oxygen saturation, microarousal index, arousal index, sleep efficiency (SE), and sleep latency (SOL), which show greater impairment in patient with COMISA[2].

In 2021, Sweetman and colleagues postulated the existence of a bidirectional causal relationship between the two disorders based on shared pathophysiology. In this model, hyperarousal caused by insomnia would heighten the perception of apneas, while OSA-related disruptions could exacerbate insomnia symptoms. Both conditions would also recruit overlapping neurobiological pathways involving the autonomic nervous system, inflammation, and arousal[1], offering potential pathophysiological overlaps. Building on this hypothesis, we hypothesize that COMISA is not merely the sum of insomnia and OSA but genuinely synergize adverse health consequences. To test this, we explore statistical interactions between insomnia and OSA on their common comorbidities, sleep symptoms, and PSG parameters in a large multi-center clinical sample.

## Methods

### Population

From 2004 to 2019, SleepMed (now BioSerenity) sleep laboratories collected data from a total of 540,941 patients who came for sleep assessment in 240 clinics across 30 states of the United States. Demographic information, medical history, and sleep symptoms were collected using a unified questionnaire. All patients provided informed consent for use of their data. The study protocol was reviewed and approved by the Advarra and Stanford institutional review boards. The details of the anonymized data set used here have been described in previous research[31].

This study includes patients whose electrophysiological assessment was a baseline PSG and who had not received a split-night PSG. Patients with PSG-derived total sleep time (TST) less than 60 min and/or missing data were excluded from the analysis. The total sample was divided into four mutually exclusive groups: 1) control (neither insomnia nor OSA), 2) insomnia alone, 3) OSA alone, and 4) COMISA (**Figure 1**).

**Figure 1.**
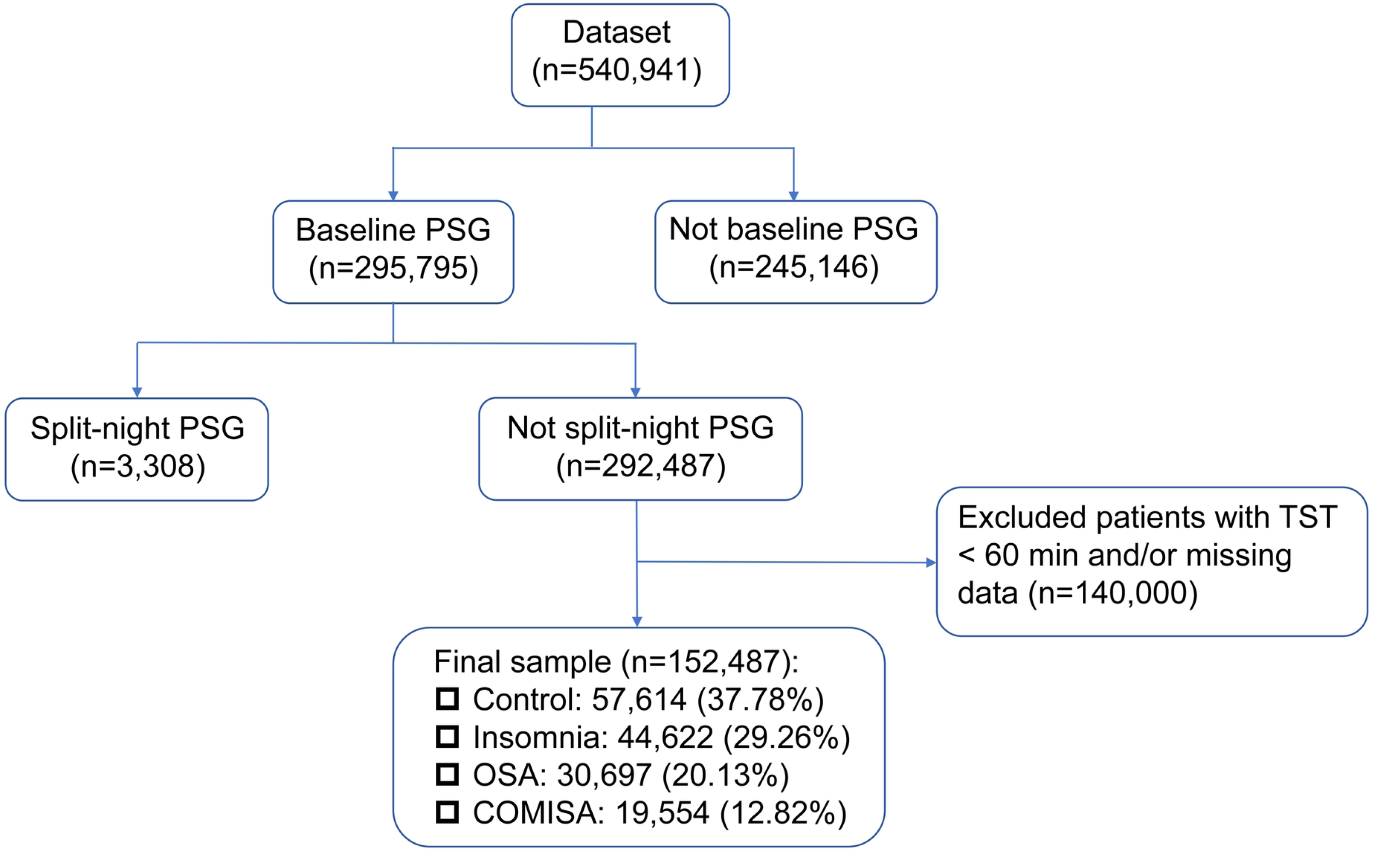
Patient Screening Flowchart. Abbreviations: OSA, obstructive sleep apnea; PSG, polysomnography; TST, total sleep time.

### Evaluation of insomnia

In the sleep assessment form, two questions were used to evaluate insomnia nocturnal symptoms: 1) having problem going to sleep at night 2) having problem waking up during the night. Three questions were used to assess daytime dysfunction: 1) waking up feeling restless 2) having problem with tiredness and 3) having problem with sleepiness. These symptoms for the past 6 months were evaluated by a five-point scale: “none” = 0, “few times” = 1, “sometimes” = 2, “often” = 3, and “always” = 4. In this study, insomnia was defined as a self-reported answer of “often” or “always” to either of the two insomnia nocturnal symptoms combined with a self-reported answer of “often” or “always” to at least one of the three daytime dysfunction questions.

### Evaluation of obstructive sleep apnea

All patients completed an overnight PSG monitoring. The recorded signals included: electroencephalograph, chin and leg electromyograms, electrocardiogram, nasal and oral breathing, chest movements, leg movements, and position. PSG parameters were extracted from sleep study reports scored by a certified sleep technician according to the American Academy of Sleep Medicine guidelines[32]. Apnea was defined as a cessation of airflow > 10s, and hypopnea was defined as a discernible reduction in breathing with a reduction in oxyhemoglobin saturation > 3%[32]. AHI refers to the total number of apneas and hypopneas per hour of sleep. In this study, we defined OSA as AHI >15 events/h, corresponding to at least moderate OSA. This threshold has been widely used in epidemiological studies because moderate-to-severe OSA is more consistently associated with adverse cardiovascular and metabolic outcomes, whereas the clinical significance of mild OSA (AHI 5–14.9 events/h) remains controversial[33].

### Evaluation of comorbidities

The comorbidities that have been reported in previous studies and are closely associated with both insomnia and OSA, including high blood pressure, diabetes, heart attack, stroke, heart enlargement, irregular heartbeat, depression, memory loss, heartburn, chronic muscular pain, headache were evaluated using self-reported questionnaire. Nocturia was defined as self-reported often or always awakening with the urgent need to urinate over the past 6 months. Subjective excessive daytime sleepiness (EDS) was defined as an Epworth Sleepiness Scale (ESS) score > 10.

### Evaluation of sleep symptoms

Common sleep symptoms related to insomnia and OSA, including falling asleep with thought racing, depressed, anxious, muscular tension, afraid of not being able to sleep, restlessness in legs and pain/discomfort were evaluated by the sleep-related questionnaire. These symptoms for the past 6 months were evaluated using a five-point scale: “none”, “few times”, “sometimes”, “often”, and “always”. In this study, we transformed the above symptoms into binary variables by converting a response of “often” or “always” to 1 and the remaining responses to 0. Weekday and weekend naps were also evaluated.

### Evaluation of PSG parameters

For analysis of PSG parameters, the following parameters were included: TST, SE, wake after sleep onset (WASO), SOL, REM onset latency (ROL), percentage of sleep stages (N1%, N2%, N3%, and REM%), AHI, oxygen desaturation index (ODI), respiratory disturbance index (RDI), minimum SpO_2_, total arousal index (total AI), spontaneous AI, respiratory AI, periodic limb movement–related AI (PLMAI), micro AI, awakening index and periodic leg movement index (PLMI).

### Statistical analysis

Patients’ demographic, sleep and clinical characteristics were compared between groups using Fisher’s exact test or the Mann–Whitney U test, as appropriate. Because all continuous variables were non-normally distributed, these are presented as median (25th percentile, 75th percentile). Modified Poisson regression with robust error variances[34] was conducted to evaluate the interactions between insomnia and OSA on binary outcomes, including comorbidities and sleep symptoms. Models were adjusted for age, gender, BMI, race, tobacco use, alcohol consumption, caffeinated beverage use, and daily use of sleep aids. Multiplicative interaction was assessed by including the interaction term Insomnia×OSA in models. We also assessed the main effects of insomnia and OSA by not including interaction term Insomnia×OSA in the model. If a multiplicative interaction was significant, simple slope analysis was conducted. Additive interaction was examined using risk ratio (RR) estimates from the same Poisson models[35]. The relative excess risk due to interaction (RERI), attributable proportion (AP), and synergy index (S), were calculated based on the following formulas:

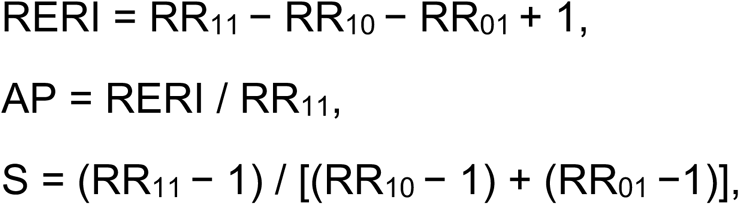

where RR_10_, RR_01_, and RR_11_ represent the RRs for exposure to insomnia alone, OSA alone, and both exposures, respectively.

Because linear regression provides estimates on the absolute scale[35], linear regression models were used to evaluate additive interactions between insomnia and OSA on PSG parameters. Additive interaction was assessed by including the interaction term Insomnia×OSA in the models. When assessing the main effects of insomnia and OSA, the interaction term was omitted. All models were adjusted for age, gender, BMI, race, tobacco use, alcohol consumption, caffeinated beverage use, and daily use of sleep aids. For outcomes showing a significant additive interaction, simple slope analysis was performed.

To account for multiple testing across all outcomes, multiple comparisons were addressed by controlling the false discovery rate (FDR). Outcomes were grouped into comorbidities, sleep symptoms and PSG parameters families, and p values of main effects and interaction terms within each family were adjusted using the Benjamini-Hochberg method[36].

Three-way interaction analyses between insomnia, OSA, and gender (male) were conducted for outcomes with significant insomnia×OSA interactions. The models were adjusted for the same set of covariates.

All analyses were completed using R (version 4.5.2), with a statistical significance of p < 0.05.

## Results

A total of 152,487 patients of median age 53 (41, 64) years, 48.9% female, including 57,614 controls, 44,622 patients with insomnia alone, 30,697 patients with OSA alone, and 19,554 patients with COMISA, were included. The demographic, comorbidities, sleep symptoms and PSG parameters of the patients stratified by insomnia and OSA are shown in **Table 1**.

**Table 1.**
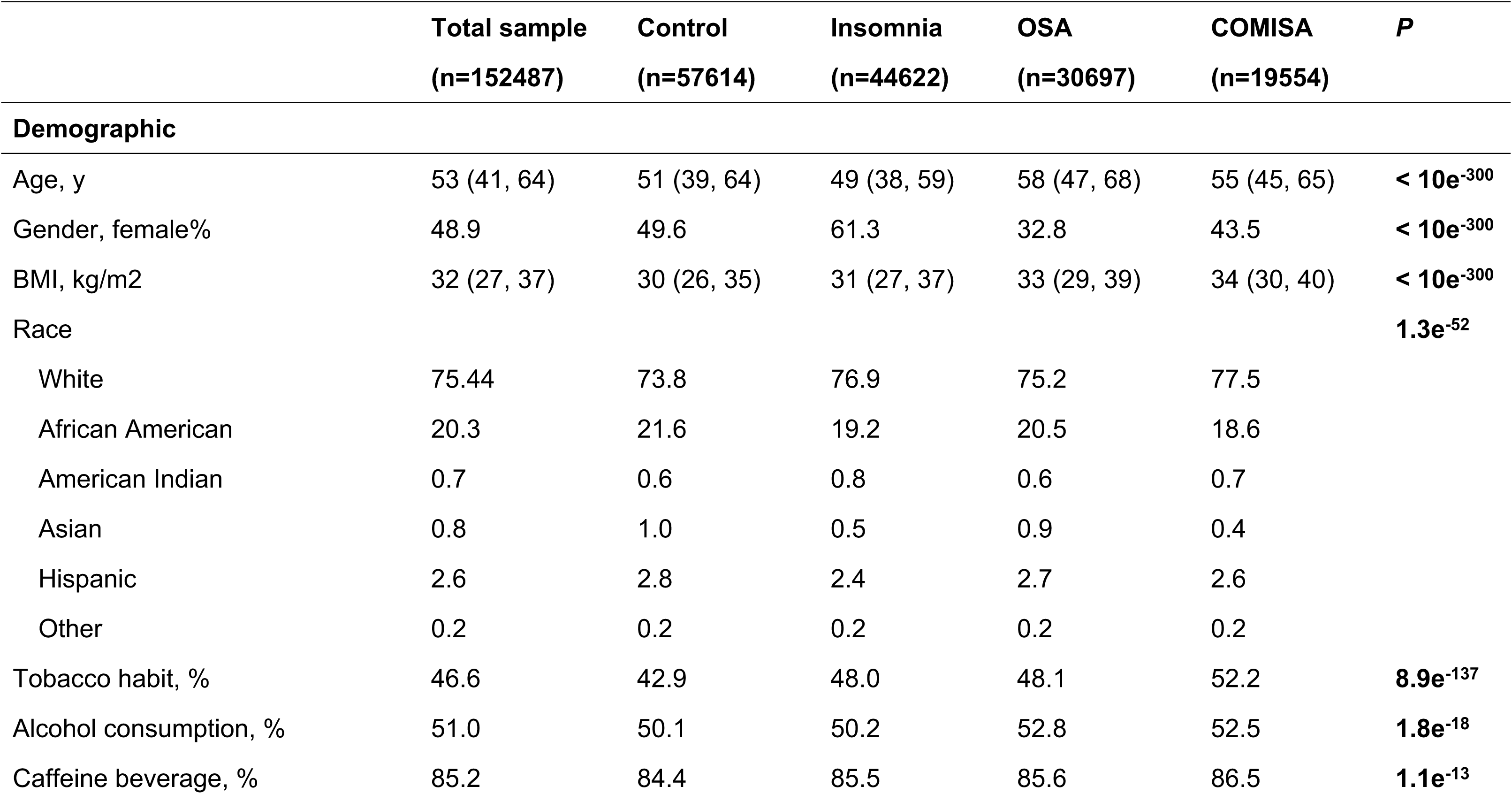
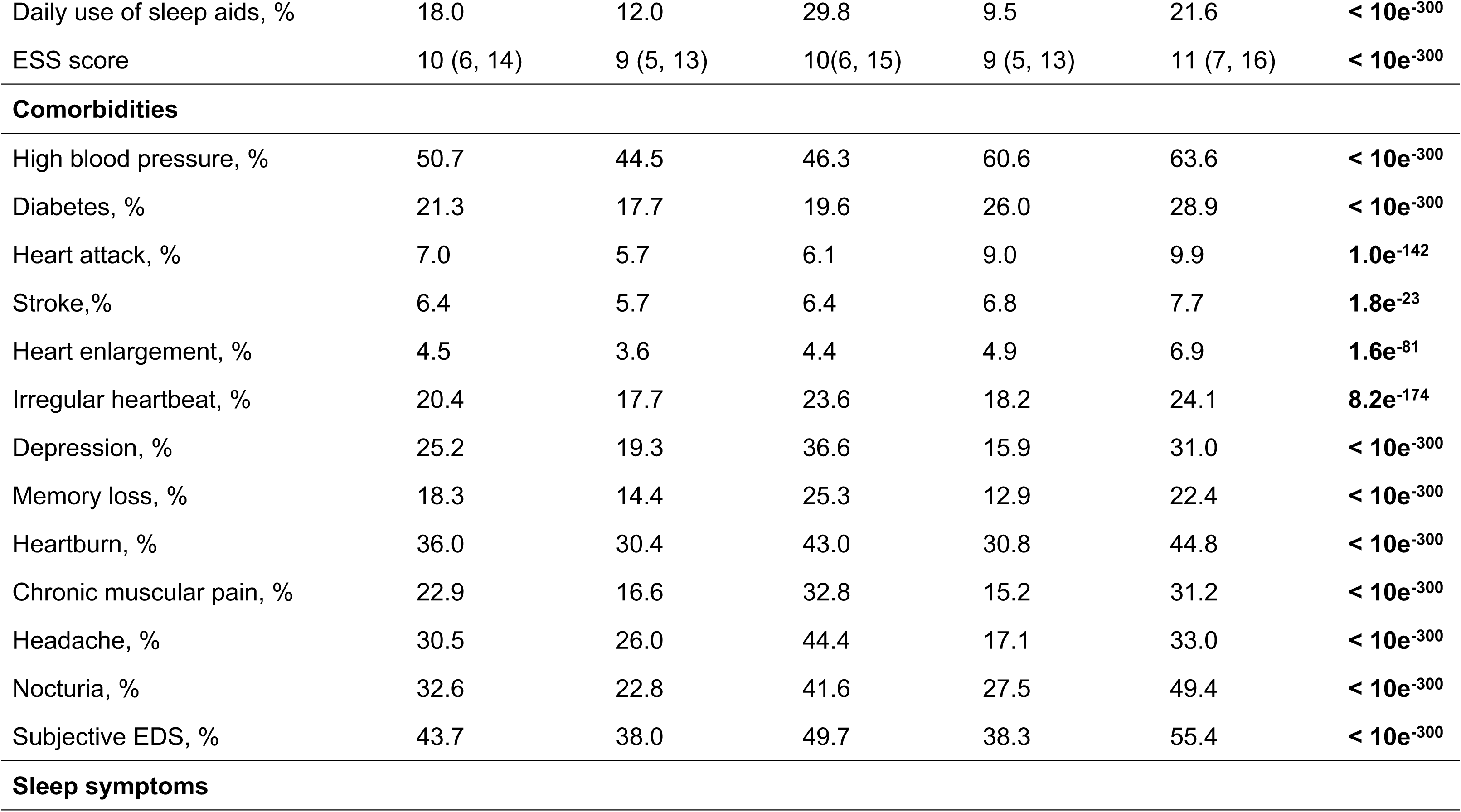
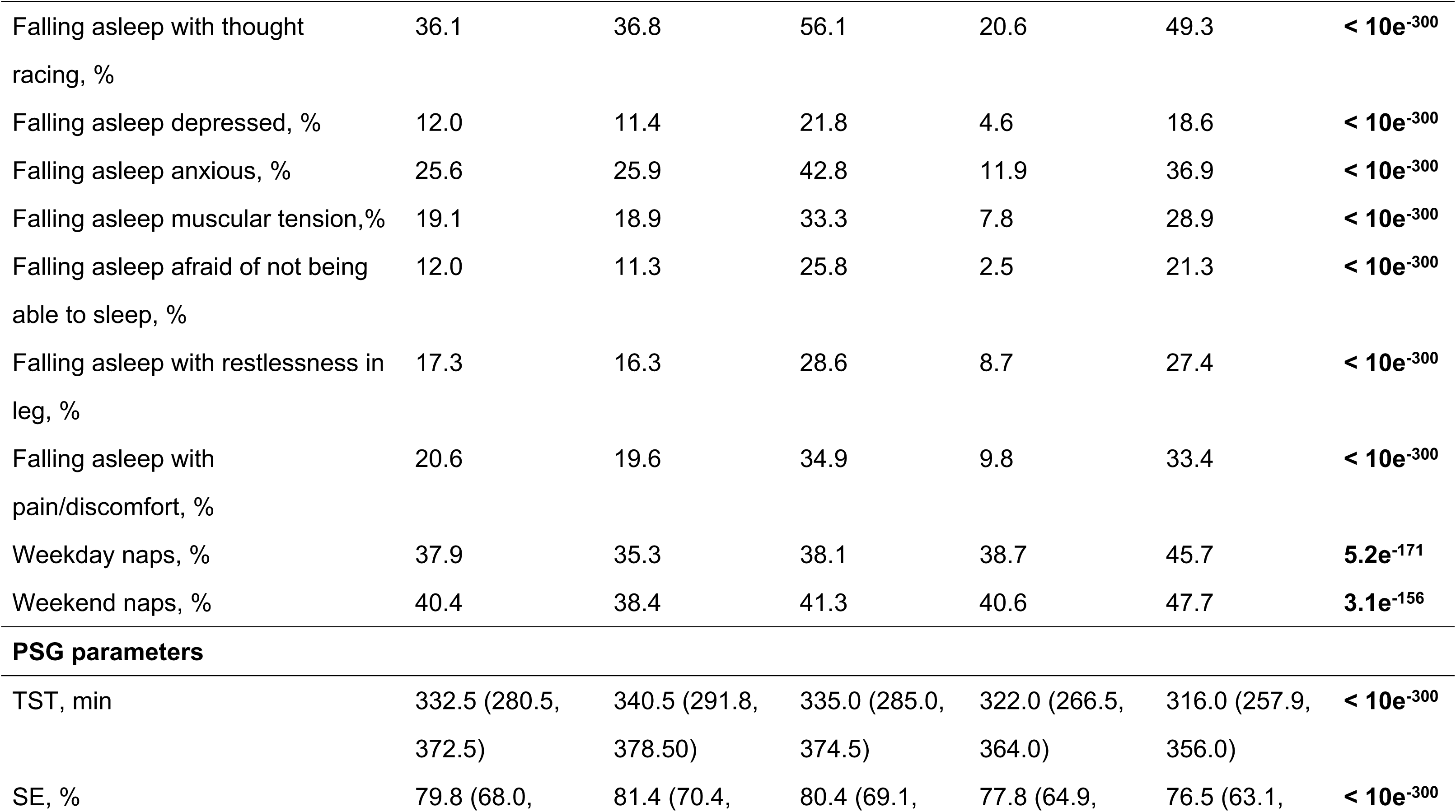
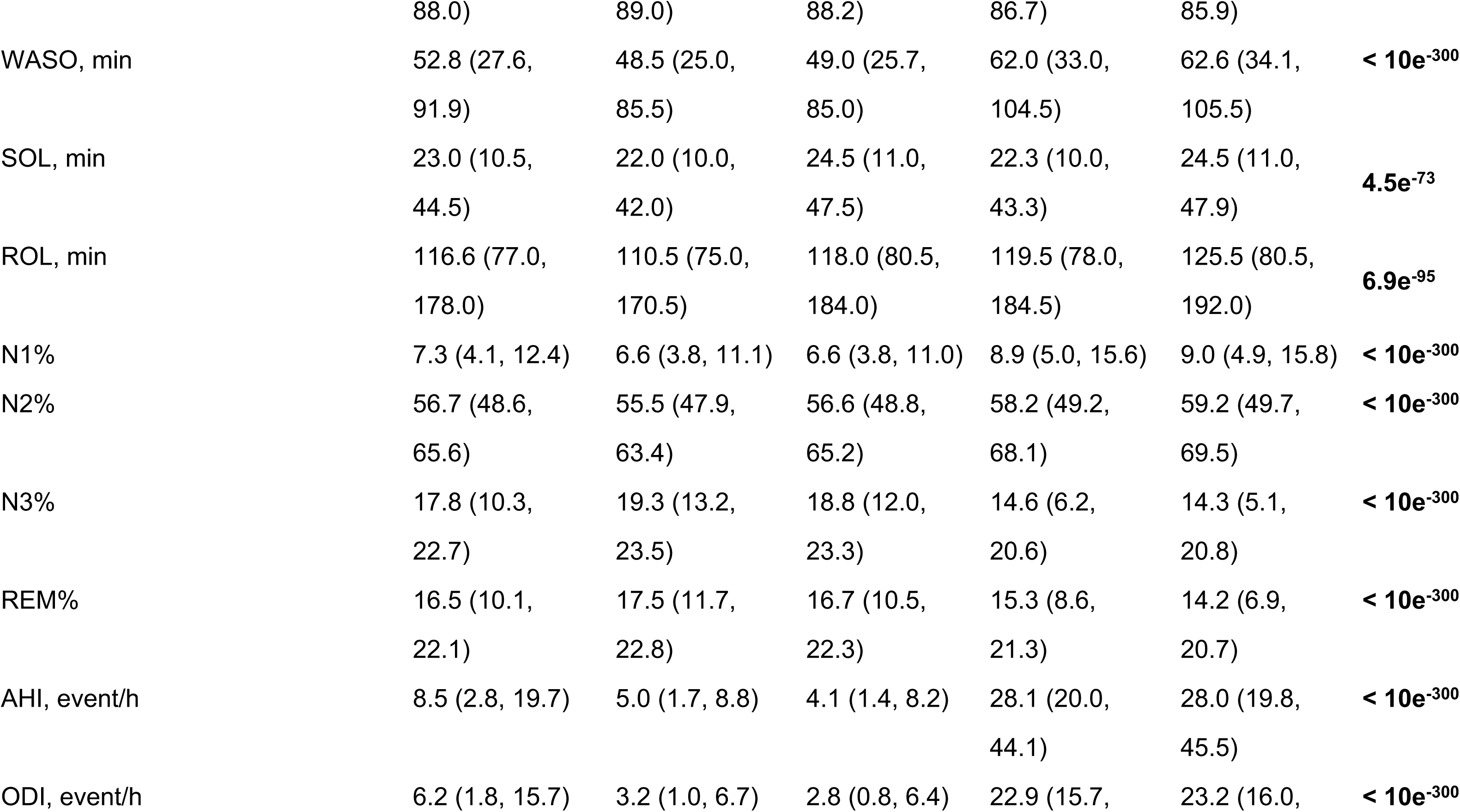
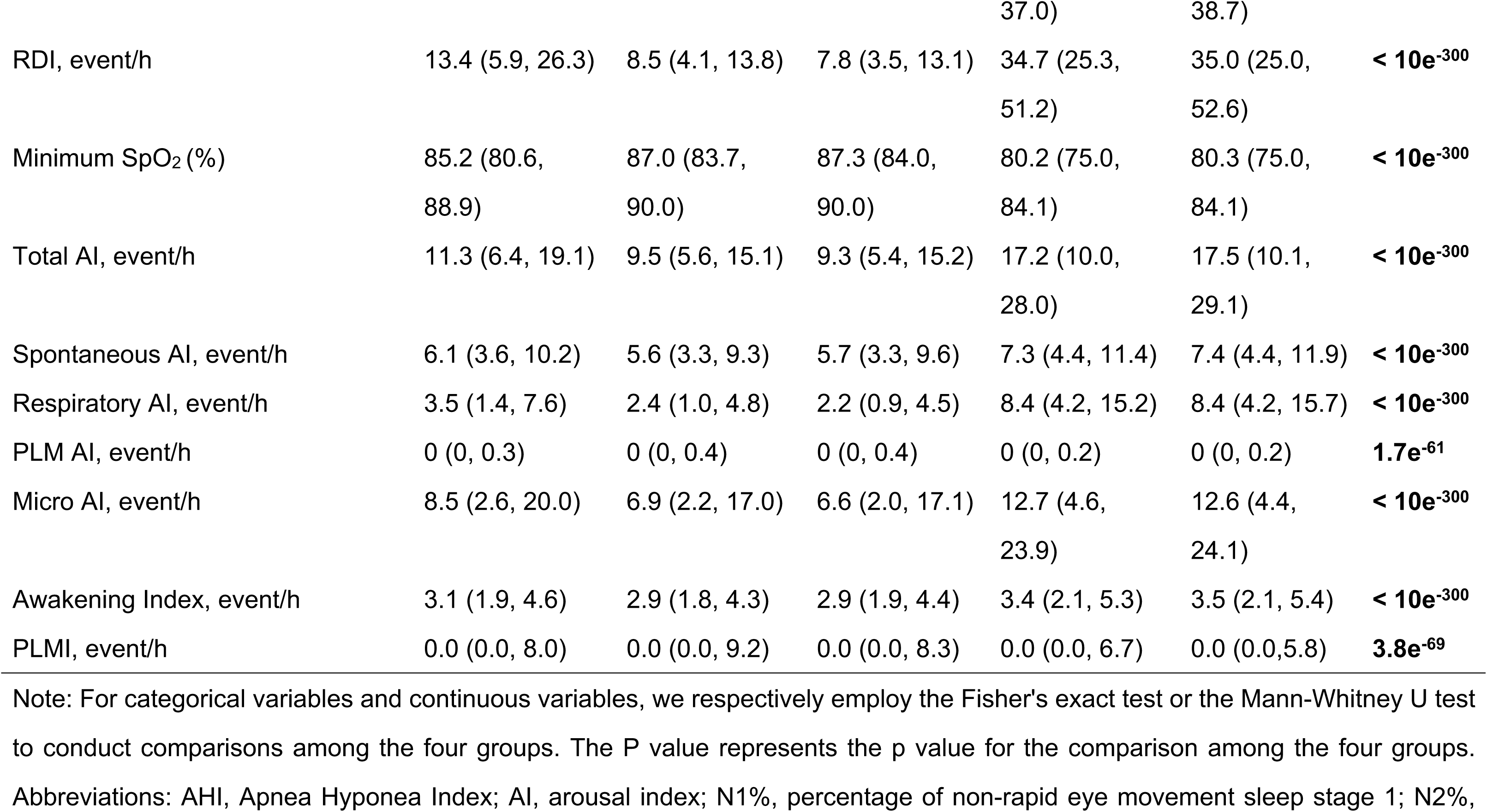
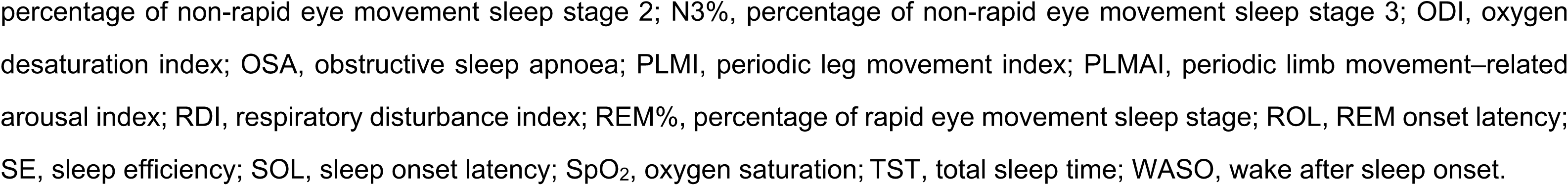
Characteristics of Patients Stratified by the Presence of Insomnia and Obstructive Sleep Apnea.

### Main effects of insomnia and OSA on comorbidities

After adjustment for confounders, insomnia was associated with increased risk of high blood pressure (RR=1.069, 95%CI=[1.059, 1.079], adjusted p=2.7e^- 43^), diabetes (RR=1.130, 95%CI=[1.108, 1.151], adjusted p=6.4e^-36^), heart attack (RR=1.287, 95%CI=[1.240, 1.335], adjusted p=8.0e^-41^), stroke (RR=1.203, 95%CI=[1.156, 1.251], adjusted p=4.7e^-20^), heart enlargement (RR=1.345, 95%CI=[1.283, 1.410], adjusted p=1.1e^-34^), irregular heartbeat (RR=1.320, 95%CI=[1.294, 1.347], adjusted p=2.0e^-160^), depression (RR=1.527, 95%CI=[1.500, 1.555], adjusted p < 10e^-300^), memory loss (RR=1.613, 95%CI=[1.578, 1.649], adjusted p < 10e^-300^), heartburn (RR=1.337, 95%CI=[1.319, 1.356], adjusted p < 10e^-300^), chronic muscular pain (RR=1.755, 95%CI=[1.722, 1.789], adjusted p < 10e^-300^), headache (RR=1.546, 95%CI=[1.523, 1.570], adjusted p < 10e^-300^), nocturia (RR=1.794, 95%CI=[1.767, 1.820], adjusted p < 10e^-300^) and subjective EDS (RR=1.376, 95%CI=[1.360, 1.392], adjusted p < 10e^-300^) (**Figure 2.A**).

**Figure 2.**
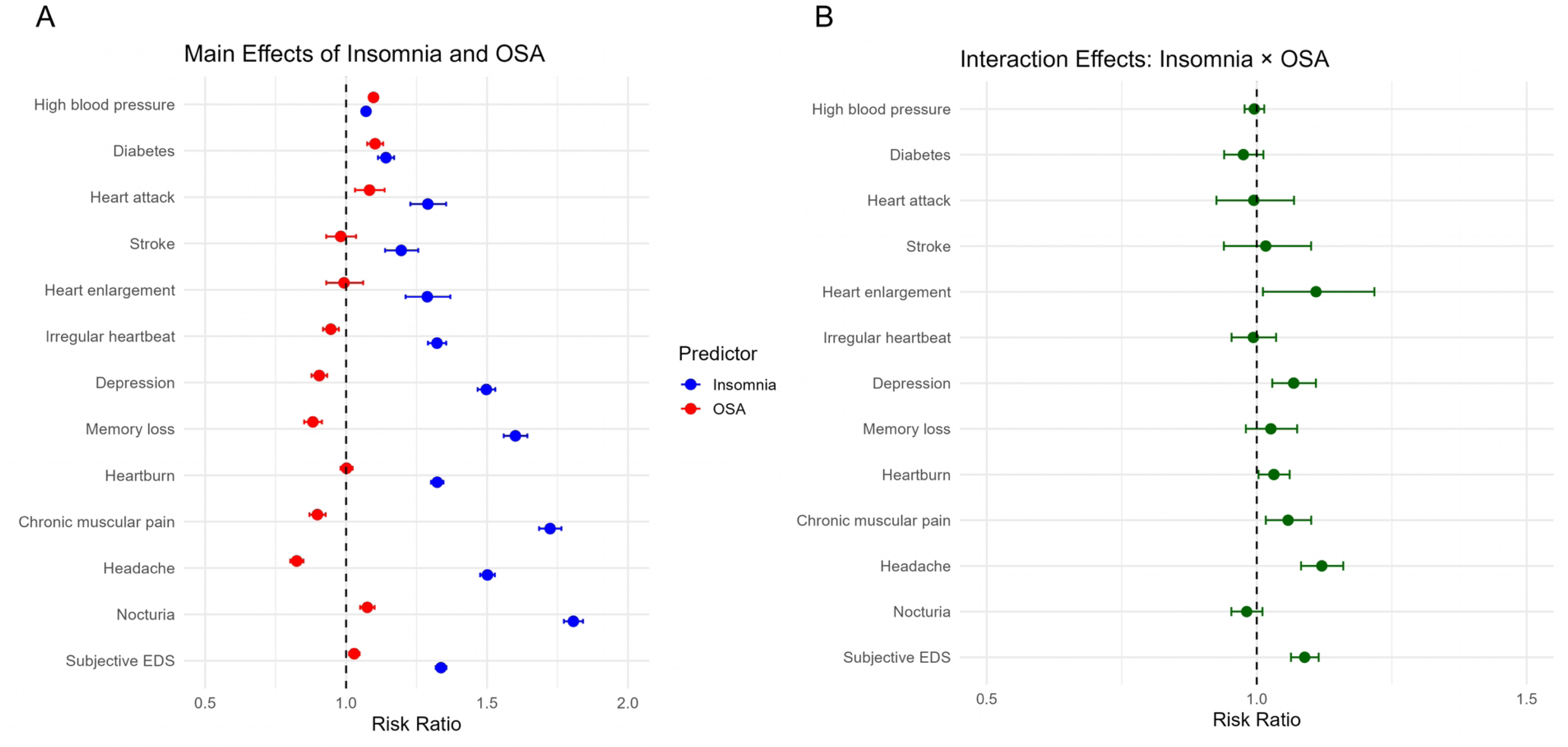
Forest plot of the main effects and interactions between insomnia and OSA on comorbidities. Note: A. Forest plot of the main effects of insomnia and OSA on comorbidities; B. Forest plot of the interactions between insomnia and OSA on comorbidities. Abbreviations: EDS, excessive daytime sleepiness; OSA, obstructive sleep apnea.

OSA was associated with increased risk of high blood pressure (RR=1.095, 95%CI=[1.085, 1.106], adjusted p=1.6e^-76^), diabetes (RR=1.091, 95%CI=[1.070, 1.113], adjusted p=4.9e^-18^), heart attacks (RR=1.081, 95%CI=[1.041, 1.122], adjusted p=6.9e^-5^), heartburn (RR=1.017, 95%CI=[1.002, 1.033], adjusted p=0.031), nocturia (RR=1.064, 95%CI=[1.048, 1.080], adjusted p=1.7e^-15^) and subjective EDS (RR=1.072, 95%CI=[1.059, 1.086], adjusted p=1.6e^-26^). However, OSA was protective against irregular heartbeat (RR=0.943, 95%CI=[0.923, 0.964], adjusted p=1.9e^-7^), depression (RR=0.939, 95%CI=[0.921, 0.958], adjusted p=6.3e^-10^), memory loss (RR=0.895, 95%CI=[0.873, 0.917], adjusted p=7.0e^-19^), chronic muscular pain (RR=0.928, 95%CI=[0.909, 0.947], adjusted p=9.6e^-13^) and headache (RR=0.878, 95%CI=[0.862, 0.894], adjusted p=4.0e^-43^) (**Figure 2.A**).

### Interactions between insomnia and OSA on comorbidities

On the relative risk scale (**Figure 2.B, Table S1**), after adjustment for confounders, insomnia and OSA showed significant positive multiplicative interactions for depression (RR=1.068, 95%CI=[1.029, 1.110], adjusted p=0.003), chronic muscular pain (RR=1.058, 95%CI=[1.017, 1.101], adjusted p=0.017), headache (RR=1.121, 95%CI=[1.082, 1.160], adjusted p=1.1e^-9^) and subjective EDS (RR=1.089, 95%CI=[1.063, 1.115], adjusted p=1.6e^-11^). However, no multiplicative interaction between them was observed for high blood pressure, diabetes, heart attack, stroke, irregular heartbeat and memory loss.

On the absolute risk scale (**Table 2**), positive additive interactions between insomnia and OSA were observed for heart enlargement (RERI=0.138, AP=0.098, S=1.492, p=0.014), heartburn (RERI=0.043, AP=0.031, S=1.131, p=0.011), depression (RERI=0.045, AP=0.031, S=1.113, p=0.011), headache (RERI=0.062, AP=0.044, S=1.189, p=0.002) and subjective EDS (RERI=0.132, AP=0.088, S=1.360, p < 10e^-300^).

**Table 2.**
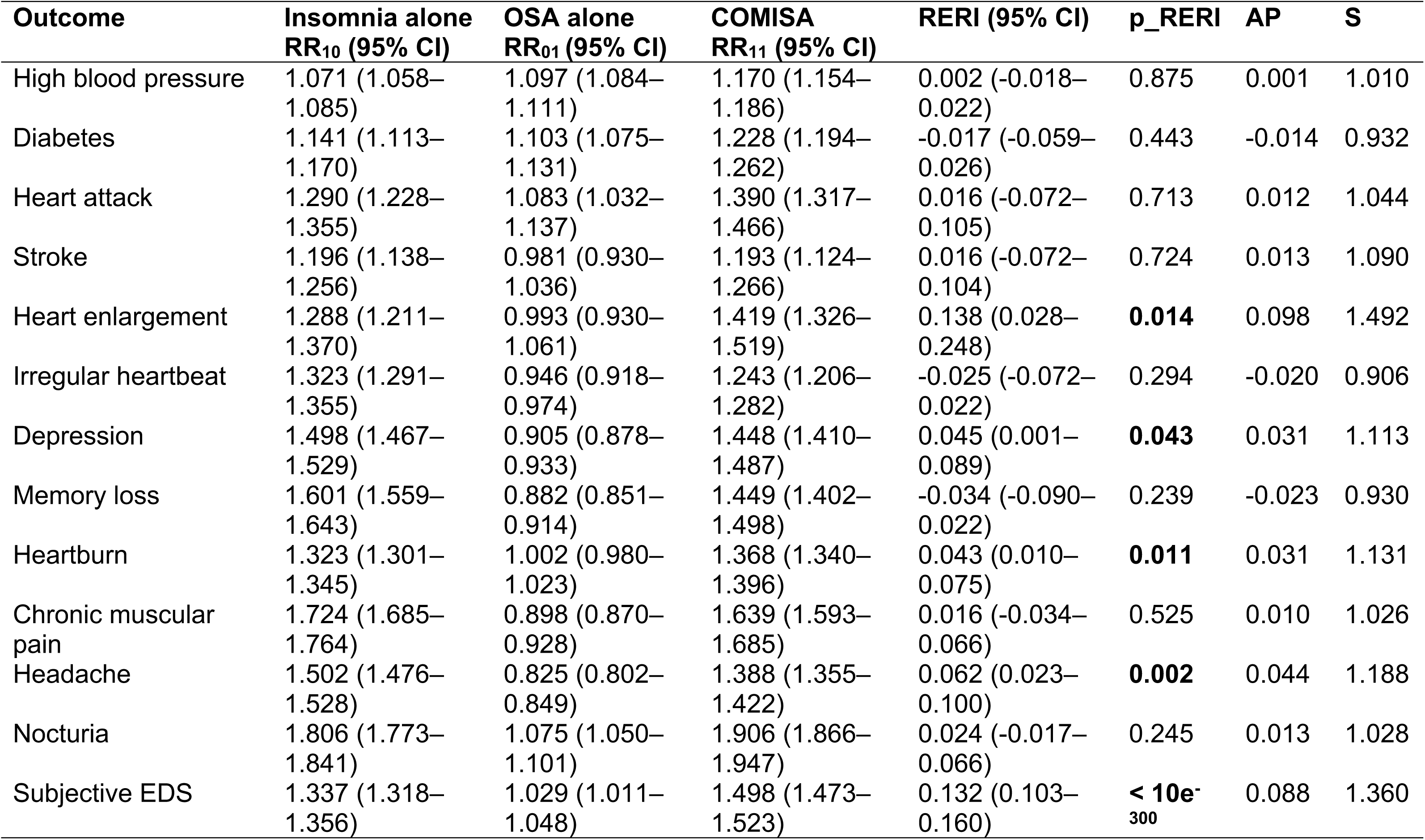
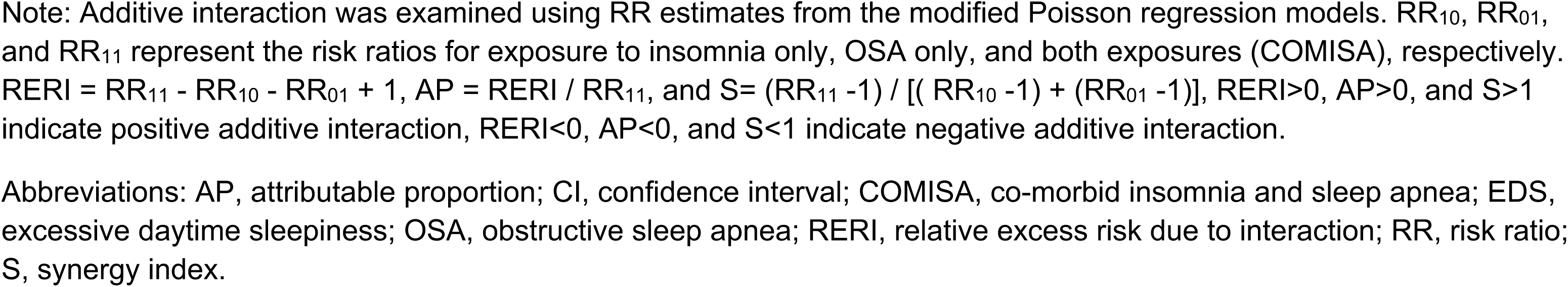
Additive Interaction between Insomnia and OSA on Comorbidities.

| **Outcome** | **Insomnia alone**<br>**RR<sub>10</sub> (95% CI)** | **OSA alone**<br>**RR<sub>01</sub> (95% CI)** | **COMISA**<br>**RR<sub>11</sub> (95% CI)** | **RERI (95% CI)** | **p_RERI** | **AP** | **S** |
| --- | --- | --- | --- | --- | --- | --- | --- |
| High blood pressure | 1.071 (1.058–1.085) | 1.097 (1.084–1.111) | 1.170 (1.154–1.186) | 0.002 (-0.018–0.022) | 0.875 | 0.001 | 1.010 |
| Diabetes | 1.141 (1.113–1.170) | 1.103 (1.075–1.131) | 1.228 (1.194–1.262) | -0.017 (-0.059–0.026) | 0.443 | -0.014 | 0.932 |
| Heart attack | 1.290 (1.228–1.355) | 1.083 (1.032–1.137) | 1.390 (1.317–1.466) | 0.016 (-0.072–0.105) | 0.713 | 0.012 | 1.044 |
| Stroke | 1.196 (1.138–1.256) | 0.981 (0.930–1.036) | 1.193 (1.124–1.266) | 0.016 (-0.072–0.104) | 0.724 | 0.013 | 1.090 |
| Heart enlargement | 1.288 (1.211–1.370) | 0.993 (0.930–1.061) | 1.419 (1.326–1.519) | 0.138 (0.028–0.248) | **0.014** | 0.098 | 1.492 |
| Irregular heartbeat | 1.323 (1.291–1.355) | 0.946 (0.918–0.974) | 1.243 (1.206–1.282) | -0.025 (-0.072–0.022) | 0.294 | -0.020 | 0.906 |
| Depression | 1.498 (1.467–1.529) | 0.905 (0.878–0.933) | 1.448 (1.410–1.487) | 0.045 (0.001–0.089) | **0.043** | 0.031 | 1.113 |
| Memory loss | 1.601 (1.559–1.643) | 0.882 (0.851–0.914) | 1.449 (1.402–1.498) | -0.034 (-0.090–0.022) | 0.239 | -0.023 | 0.930 |
| Heartburn | 1.323 (1.301–1.345) | 1.002 (0.980–1.023) | 1.368 (1.340–1.396) | 0.043 (0.010–0.075) | **0.011** | 0.031 | 1.131 |
| Chronic muscular pain | 1.724 (1.685–1.764) | 0.898 (0.870–0.928) | 1.639 (1.593–1.685) | 0.016 (-0.034–0.066) | 0.525 | 0.010 | 1.026 |
| Headache | 1.502 (1.476–1.528) | 0.825 (0.802–0.849) | 1.388 (1.355–1.422) | 0.062 (0.023–0.100) | **0.002** | 0.044 | 1.188 |
| Nocturia | 1.806 (1.773–1.841) | 1.075 (1.050–1.101) | 1.906 (1.866–1.947) | 0.024 (-0.017–0.066) | 0.245 | 0.013 | 1.028 |
| Subjective EDS | 1.337 (1.318–1.356) | 1.029 (1.011–1.048) | 1.498 (1.473–1.523) | 0.132 (0.103–0.160) | **< 10e-<sub>300</sub>** | 0.088 | 1.360 |

Note: Additive interaction was examined using RR estimates from the modified Poisson regression models. $RR_{10}$ , $RR_{01}$ , and $RR_{11}$ represent the risk ratios for exposure to insomnia only, OSA only, and both exposures (COMISA), respectively. $RERI = RR_{11} - RR_{10} - RR_{01} + 1$ , $AP = RERI / RR_{11}$ , and $S = (RR_{11} - 1) / [(RR_{10} - 1) + (RR_{01} - 1)]$ , $RERI > 0$ , $AP > 0$ , and $S > 1$ indicate positive additive interaction, $RERI < 0$ , $AP < 0$ , and $S < 1$ indicate negative additive interaction.
Abbreviations: AP, attributable proportion; CI, confidence interval; COMISA, co-morbid insomnia and sleep apnea; EDS, excessive daytime sleepiness; OSA, obstructive sleep apnea; RERI, relative excess risk due to interaction; RR, risk ratio; S, synergy index.

Simple slope analyses (**Figure S1**) showed that the associations between insomnia and those comorbidities were consistently stronger among patients with OSA than among those without OSA.

### Main effects of insomnia and OSA on sleep symptoms

As shown in **Figure 3.A**, after adjustment for confounders, not surprisingly, insomnia was associated with increased risk of falling asleep with thoughts racing (RR=2.151, 95%CI=[2.120, 2.183], adjusted p < 10e^-300^), depressed (RR=3.120, 95%CI=[3.020, 3.224], adjusted p < 10e^-300^), anxious (RR=2.470, 95%CI=[2.423, 2.519], adjusted p < 10e^-300^), muscular tension (RR=2.781, 95%CI=[2.714, 2.849], adjusted p < 10e^-300^), afraid of not being able to sleep (RR=6.623, 95%CI=[6.354, 6.904], adjusted p < 10e^-300^), restlessness in legs (RR=2.709, 95%CI=[2.642, 2.778], adjusted p < 10e^-300^) and pain/discomfort (RR=2.812, 95%CI=[2.749, 2.877], adjusted p < 10e^-300^).

**Figure 3.**
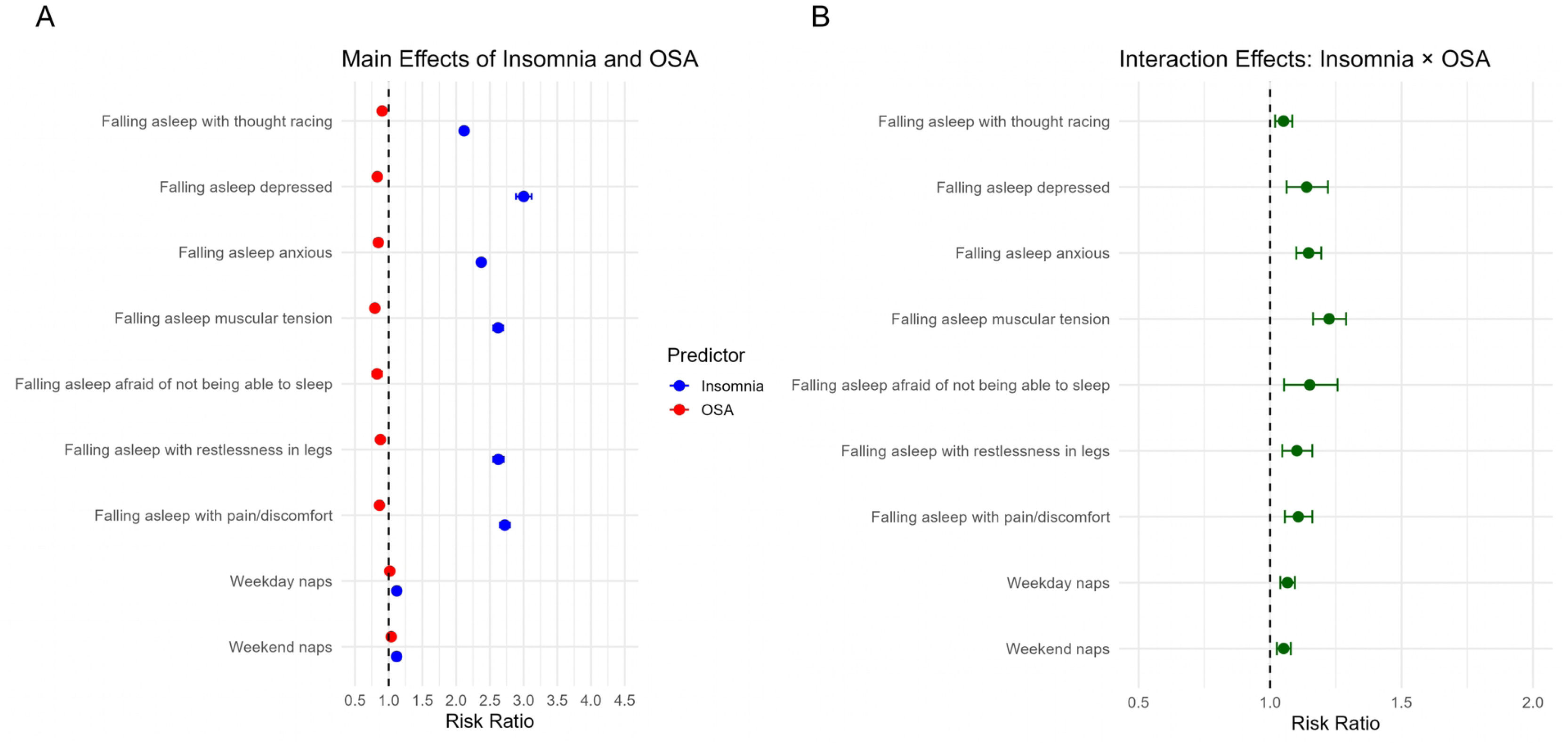
Forest plot of the main effects and interactions between insomnia and OSA on sleep symptoms. Note: A. Forest plot of the main effects of insomnia and OSA on sleep symptoms; B. Forest plot of the interactions between insomnia and OSA on sleep symptoms. Abbreviations: EDS, excessive daytime sleepiness; OSA, obstructive sleep apnea.

On contrary, OSA was protective against falling asleep with thoughts racing (RR=0.931, 95%CI=[0.917, 0.945], adjusted p=2.2e^-19^), depressed (RR=0.913, 95%CI=[0.885, 0.942], adjusted p=2.0e^-8^), anxious (RR=0.928, 95%CI=[0.910, 0.946], adjusted p=1.8e^-13^), muscular tension (RR=0.915, 95%CI=[0.894, 0.937], adjusted p=4.5e^-13^), afraid of not being able to sleep (RR=0.932, 95%CI=[0.904, 0.961], adjusted p=8.2e^-6^), restlessness in legs (RR=0.937, 95%CI=[0.914, 0.960], adjusted p=2.2e^-7^) and pain/discomfort (RR=0.927, 95%CI=[0.907, 0.947], adjusted p=1.0e^-11^).

Both insomnia and OSA were associated with weekday (insomnia: RR=1.146, 95%CI=[1.131, 1.161], adjusted p=1.2e^-92^; OSA: RR=1.046, 95%CI=[1.032, 1.061], adjusted p=1.4e^-10^) and weekend naps (insomnia: RR=1.138, 95%CI=[1.124, 1.152], adjusted p=1.9e^-90^; OSA: RR=1.061, 95%CI=[1.047, 1.076], adjusted p=1.0e^-17^).

### Interactions between insomnia and OSA on sleep symptoms

On the relative risk scale (**Figure 3.B, Table S1**), after adjustment for confounders, insomnia and OSA demonstrated strong significant positive multiplicative interactions for falling asleep with thoughts racing (RR=1.051, 95%CI=[1.019, 1.084], adjusted p=0.002), depressed (RR=1.139, 95%CI=[1.063, 1.220], adjusted p=2.9e^-4^), anxious (RR=1.146, 95%CI=[1.100, 1.195], adjusted p=3.9e^-10^), muscular tension (RR=1.225, 95%CI=[1.163, 1.289], adjusted p=1.1e^-13^), afraid of not being able to sleep (RR=1.151, 95%CI=[1.054, 1.257], adjusted p=0.002), restlessness in legs (RR=1.102, 95%CI=[1.047, 1.160], adjusted p=2.9e^-4^), pain/discomfort (RR=1.107, 95%CI=[1.057, 1.160], adjusted p=4.5e^-5^), weekday naps (RR=1.066, 95%CI=[1.039, 1.094], adjusted p=3.9e^-6^) and weekend naps (RR=1.052, 95%CI=[1.026, 1.079], adjusted p=1.3e^-4^).

On the absolute risk scale (**Table 3**), insomnia and OSA showed significant positive additive interactions on falling asleep with anxious (RERI=0.085, AP=0.037, S=1.070, p=0.004), muscular tension (RERI=0.136, AP=0.053, S=1.096, p=4.3e^-4^), weekday naps (RERI=0.078, AP=0.064, S=1.564, p=8.6e^- 8^) and weekend naps (RERI=0.065, AP=0.053, S=1.417, p=4.3e^-6^).

**Table 3.** Additive Interaction between Insomnia and OSA on Sleep Symptoms.

| **Outcome** | **Insomnia alone**<br>**RR<sub>10</sub> (95% CI)** | **OSA alone**<br>**RR<sub>01</sub> (95% CI)** | **COMISA**<br>**RR<sub>11</sub> (95% CI)** | **RERI (95% CI)** | **p_RERI** | **AP** | **S** |
| --- | --- | --- | --- | --- | --- | --- | --- |
| Falling asleep with thought racing | 2.119 (2.083, 2.155) | 0.903 (0.879, 0.927) | 2.011 (1.970, 2.053) | -0.011 (-0.052, 0.031) | 0.623 | -0.005 | 0.990 |
| Falling asleep depressed | 3.003 (2.890–3.120) | 0.831 (0.782–0.884) | 2.844 (2.720–2.973) | 0.010 (-0.104–0.123) | 0.869 | 0.003 | 1.005 |
| Falling asleep anxious | 2.374 (2.321–2.428) | 0.848 (0.818–0.879) | 2.307 (2.246–2.370) | 0.085 (0.027–0.144) | **0.004** | 0.037 | 1.070 |
| Falling asleep muscular tension | 2.623 (2.551–2.697) | 0.795 (0.760–0.832) | 2.554 (2.472–2.639) | 0.136 (0.060–0.212) | **4.3e-4** | 0.053 | 1.096 |
| Falling asleep afraid of not being able to sleep | 6.355 (6.056–6.670) | 0.828 (0.762–0.900) | 6.055 (5.745–6.383) | -0.128 (-0.339–0.084) | 0.236 | -0.021 | 0.975 |
| Falling asleep with restlessness in legs | 2.627 (2.550–2.706) | 0.878 (0.839–0.918) | 2.540 (2.454–2.630) | 0.036 (-0.045–0.117) | 0.386 | 0.014 | 1.024 |
| Falling asleep with pain/discomfort | 2.722 (2.649–2.797) | 0.864 (0.829–0.901) | 2.606 (2.526–2.687) | 0.019 (-0.053–0.091) | 0.606 | 0.007 | 1.012 |
| Weekday naps | 1.120 (1.102–1.139) | 1.018 (0.999–1.036) | 1.215 (1.192–1.239) | 0.078 (0.049–0.106) | **8.6e-8** | 0.064 | 1.564 |
| Weekend naps | 1.118 (1.100–1.135) | 1.038 (1.020–1.057) | 1.220 (1.198–1.243) | 0.065 (0.037–0.092) | **4.3e-6** | 0.053 | 1.028 |

Note: Additive interaction was examined using RR estimates from the modified Poisson regression models. RR<sub>10</sub>, RR<sub>01</sub>, and RR<sub>11</sub> represent the risk ratios for exposure to insomnia alone, OSA alone, and both exposures (COMISA), respectively. $RERI = RR_{11} - RR_{10} - RR_{01} + 1$ , $AP = RERI / RR_{11}$ , and $S = (RR_{11} - 1) / [(RR_{10} - 1) + (RR_{01} - 1)]$ , $RERI > 0$ , $AP > 0$ , and $S > 1$ indicate positive additive interaction, $RERI < 0$ , $AP < 0$ , and $S < 1$ indicate negative additive interaction.
Abbreviations: AP, attributable proportion; CI, confidence interval; COMISA, co-morbid insomnia and sleep apnea; OSA, obstructive sleep apnea; RERI, relative excess risk due to interaction; RR, risk ratio; S, synergy index.

Simple slope analyses (**Figure S2**) showed that the associations between insomnia and those pre-sleep symptoms were consistently stronger among patients with OSA than among those without OSA.

### Main effects of insomnia and OSA on PSG parameters

As shown in **Figure 4.A** and **Table S2**, after adjustment for confounders, insomnia was associated with decreased TST (β=-8.895, adjusted p=3.6e^-121^), SE (β=-1.873,adjusted p=1.7e^-118^), N3% (β=-0.481, adjusted p=1.7e^-20^) and REM% (β=-0.463, adjusted p=5.1e^-23^), and increased WASO (β=4.647, adjusted p=2.0e^-68^), SOL (β=2.834, adjusted p=2.4e^-49^), ROL (β=1.231, adjusted p=0.002), N1% (β=0.398, adjusted p=5.7e^-16^), N2% (β=0.551, adjusted p=3.2e^-14^), AHI (β=0.305, adjusted p=7.4e^-6^), ODI (β=0.342, adjusted p=1.6e^-7^), RDI (β=0.359, adjusted p=7.1e^-7^), minimum SpO_2_ (β=0.203, adjusted p=1.2e^-10^), total AI (β=0.676, adjusted p=1.8e^-27^), spontaneous AI (β=0.327, adjusted p=1.4e^-20^), respiratory AI (β=0.278, adjusted p=6.1e^-13^), PLM AI (β=0.074, adjusted p=6.5e^-8^), micro AI (β=0.294, adjusted p=0.014), awakening index (β=0.204, adjusted p=3.6e^-36^) and PLMI (β=0.365, adjusted p=7.6e^-4^).

**Figure 4.**
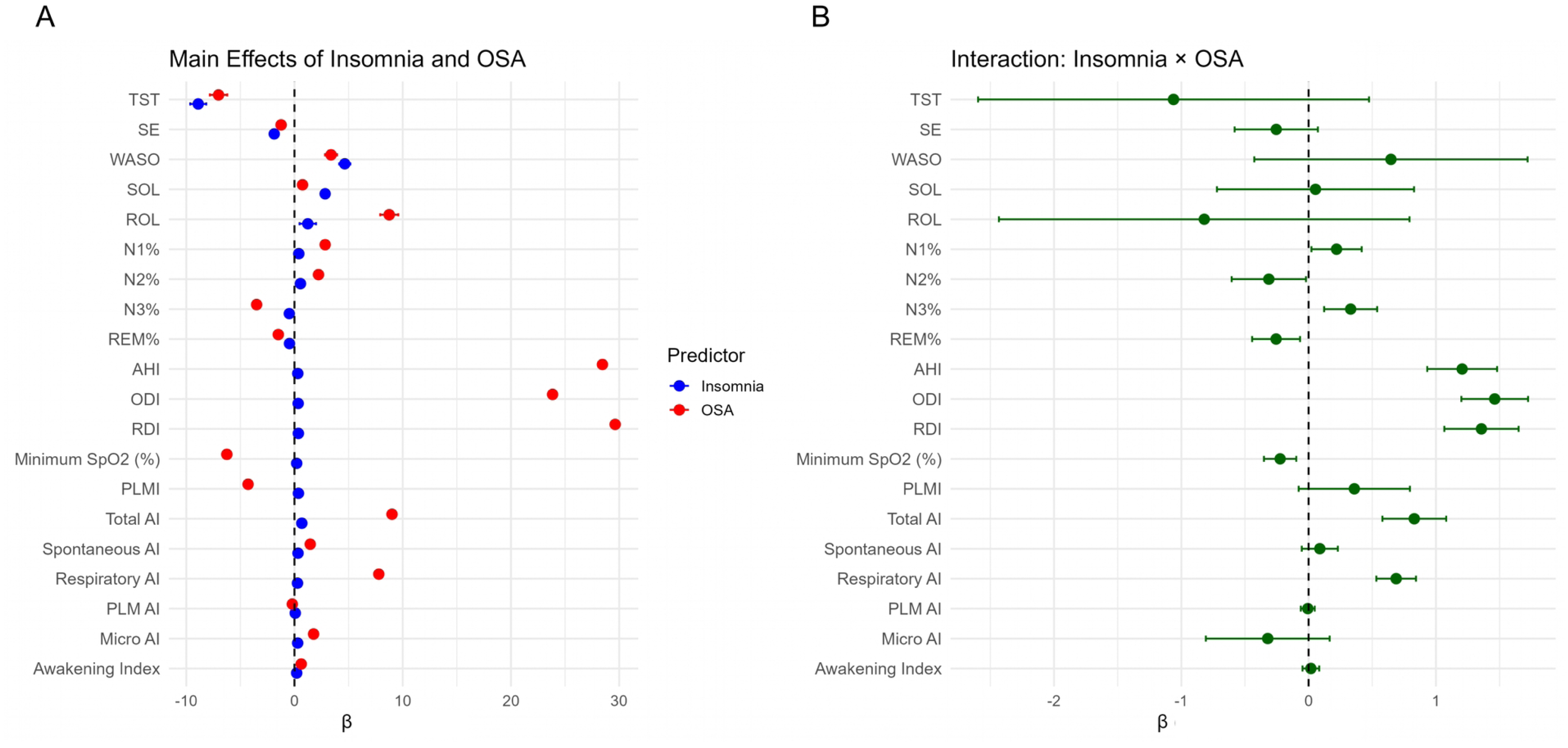
Forest plot of the main effects and interactions between insomnia and OSA on PSG parameters. Note: A. Forest plot of the main effects of insomnia and OSA on PSG parameters; B. Forest plot of the interactions between insomnia and OSA on PSG parameters. Abbreviations: AHI, Apnea Hypopnea Index; AI, arousal index; N1%, percentage of non-rapid eye movement sleep stage 1; N2%, percentage of non-rapid eye movement sleep stage 2; N3%, percentage of non-rapid eye movement sleep stage 3; ODI, oxygen desaturation index; OSA, obstructive sleep apnea; PLMI, periodic leg movement index; PLMAI, periodic limb movement–related arousal index; PSG, polysomnography; RDI, respiratory disturbance index; REM%, percentage of rapid eye movement sleep stage; ROL, REM onset latency; SE, sleep efficiency; SOL, sleep onset latency; SpO_2_, oxygen saturation; TST, total sleep time; WASO, wake after sleep onset.

OSA was associated with decreased TST (β=-7.018, adjusted p=1.3e^-65^), SE (β=-1.237, adjusted p=1.9e^-45^), N3% (β=-3.500, adjusted p < 10e^-300^), REM% (β=-1.495, adjusted p=2.2e^-193^), minimum SpO_2_ (β=-6.251, adjusted p < 10e^-300^), PLMI (β=-4.287, adjusted p=1.8e^-294^) and PLM AI (β=-0.203, adjusted p=9.4e^-44^), and increased WASO (β=3.378, adjusted p=5.0e^-32^), SOL (β=0.742, adjusted p=3.3e^-4^), ROL (β=8.755, adjusted p=1.6e^-93^), N1% (β=2.836, adjusted p < 10e^-300^), N2% (β=2.220, adjusted p=5.1e^-178^), AHI (β=28.459, adjusted p < 10e^-300^), ODI (β=23.848, adjusted p < 10e^-300^), RDI (β=29.635, adjusted p < 10e^-300^), total AI (β=9.012, adjusted p < 10e^-300^), spontaneous AI (β=1.456, adjusted p=3.5e^-322^), respiratory AI (β=7.793, adjusted p < 10e^-300^), micro AI (β=1.766, adjusted p=4.4e^-42^) and awakening index (β=0.621, adjusted p=3.6e^-274^) (**Figure 4.A, Table S2**).

### Interactions between insomnia and OSA on PSG parameters

After adjustment for confounders, insomnia and OSA demonstrated significant positive additive interactions for PSG parameters related to respiratory disturbance (**Figure 4.B, Table S2**), including AHI (β=1.207, adjusted p=3.4e^-17^), ODI (β=1.462, adjusted p=1.7e^-26^), RDI (β=1.358, adjusted p=7.3e^-19^), total AI (β=0.831, adjusted p=3.3e^-10^) and respiratory AI (β=0.688, adjusted p=2.9e^-17^). Meanwhile, insomnia and OSA demonstrated significant negative additive interactions for minimum SpO_2_ (β=-0.223, adjusted p=0.002), REM% (β=-0.255, adjusted p=0.020). Surprisingly, insomnia and OSA demonstrated significant positive additive interactions for N3% (β=0.331, adjusted p=0.005). However, no interaction between them was observed for TST, SE, WASO, SOL, ROL, N1%, PLMI, spontaneous AI, PLM AI, micro AI and awakening index.

Simple slope analyses (**Figure S3**) showed that OSA was associated with more severe respiratory disturbance (higher AHI, ODI, RDI, total AI and respiratory AI) and greater sleep disruption (lower REM%) among patients with insomnia than among those without insomnia.

### Gender difference on interactions between insomnia and OSA

For comorbidities and sleep symptoms, after adjustment for confounders, the three-way interaction term between insomnia, OSA, and gender (male) was not statistically significant for heart enlargement, depression, heartburn, chronic muscular pain, headache, subjective EDS, pre-sleep symptoms and daytime napping (**Figure S4.A**).

For PSG parameters, after adjustment for confounders, insomnia, OSA and gender (male) had positive additive interactions on AHI (β=1.347, adjusted p=2.0e^-5^), ODI (β=1.101, adjusted p=1.6e^-4^) and RDI (β=1.233, adjusted p=1.6e^-4^), and negative additive interactions on REM% (β=-0.491, adjusted p=0.025) (**Figure S4.B, Table S3**). Gender-stratified simple slope analyses showed that in males, the adverse effects of insomnia and OSA on AHI, ODI, RDI and REM% were substantially amplified when the other condition was present (**Figure S5**). By contrast, women showed minimal differences across strata of the other condition, with considerably weaker slopes for both insomnia and OSA.

## Discussion

Based on a large-scale, multi-center clinical sample, we found that both insomnia and OSA were independently associated with adverse cardiometabolic diseases, nocturia, subjective EDS and poor objective sleep quality. In contrast to OSA, insomnia was also associated with increased depression, pre-sleep cognitive-emotional and somatic hyperarousal. Most importantly, we provide for the first time evidence that insomnia and OSA truly interact to exacerbate a broad spectrum of clinical outcomes, including depression, chronic muscular pain, headache, subjective EDS, pre-sleep cognitive-emotional and somatic hyperarousal, physiological respiratory disturbances, and sleep disruption. These findings indicate that insomnia and OSA do not merely coexist but potentiate each other’s adverse health effects.

Consistent with previous findings[37–40], we found that both insomnia and OSA were independently associated with high blood pressure, diabetes and heart attack. Interestingly however, we found no interactions between them for those comorbidities. Rather, our findings suggest coexistence of insomnia and OSA is associated with excess risk for heart enlargement (presumably reflecting hypertrophy). Specifically, when insomnia and OSA coexist (COMISA), 10% of the risk of heart enlargement (AP=0.098) was caused by the interaction of the two conditions. Consistent with our findings, several cohort studies suggested that COMISA is associated with a markedly increased risk of major adverse cardiovascular events, including myocardial infarction, congestive heart failure, and stroke^[24, 41, 42]^. However, when interpreting the relationship between insomnia and OSA and their effects on heart enlargement, we should be cautious. Indeed, the multiplicative interaction showed only marginal statistical significance after FDR correction (adjusted p=0.062, Table S1).

In this study, we also found that insomnia and OSA have positive multiplicative and/or additive interactions on depression, chronic muscular pain and headache. Specifically, when insomnia and OSA coexist (COMISA), 3% of the risk of depression (AP=0.031), 4% headache (AP=0.044) was caused by the interaction of the two conditions. Our findings align with several previous studies[26–28, 43]. For example, a clinical study suggested that men with COMISA or insomnia alone had significantly higher levels of pre-sleep arousal, anxiety, and depression than the OSA group[43]. Another population-based study found that depression prevalence and symptom scale scores were highest in men with COMISA than men with insomnia or OSA alone[27]. In the present study, depression was highly prevalent among both individuals with insomnia alone (36.6%) and those with COMISA (31.0%). The slightly higher prevalence observed in the insomnia-alone group may be partly explained by the greater proportion of women in this group compared with the COMISA group (61.0% vs. 43.5%). Notably, our findings extend previous observations by demonstrating a significant biological interaction between insomnia and OSA on depression risk. By contrast, evidence regarding chronic muscular pain and headache in COMISA remains scarce. Our observation of a significant interaction between insomnia and OSA on chronic muscular pain and headache may provide novel epidemiological evidence supporting a synergistic contribution of insomnia and OSA to pain sensitization.

However, in our sample, in contrast to insomnia, the main effect of OSA on those comorbidities was “protective”, which is inconsistent with several but not all previous studies[44–46]. Although this is surprising, it could reflect the fact other studies never study OSA in the absence (or controlling for) of insomnia. Small sample size for most other studies could be another explanation as are the variable definitions of OSA (aka hypopnea definition and AHI cut offs). The large size of our dataset and the strong significance of these effects is however notable and raise doubt regarding common knowledge suggesting OSA exacerbates depression, chronic muscular pain and headaches, at least when subjectively reported.

There is solid evidence that both insomnia and OSA independently contribute to subjective EDS, or that subjective EDS is a common daytime impairment associated with both insomnia and OSA[47–49]. In this study, we found that both insomnia and OSA increased risk of subjective EDS as measured with the ESS. Furthermore, we found that the coexistence of insomnia and OSA confers an excess risk for high ESS, that exceeds the sum of individual effects. Specifically, in COMISA, 9% of the risk of subjective EDS (AP=0.088) is caused by the interaction itself of the two conditions. Consistent with these findings, a previous study suggested that in patients with OSA, insomnia severity (measured by the Insomnia Severity Index, ISI) correlates more strongly with ESS than any PSG parameters; in multivariable models, insomnia-related daily functioning interference (ISI item 7) was the only independent predictor of ESS[50].

The underlying pathological mechanism of the interactions between insomnia and OSA on those comorbidities remains unclear. The coexistence of insomnia and OSA may give rise to a synergistic pathophysiological phenotype characterized by the amplification of both biological disturbances (sleep fragmentation and intermittent hypoxia) and psychological hyperarousal. A synergistic biological interplay between overlapping sleep fragmentation in insomnia and intermittent hypoxia in OSA has been suggested[51, 52]. As expected, having OSA per our definition is associated with more severe respiratory disturbances. More interestingly, it is also associated with a greater sleep disruption, reflected by higher arousal, lower REM sleep, which could interact with typical insomnia findings of decrease sleep efficiency. Notably, these associations were consistently stronger among patients with coexisting insomnia than among those without insomnia. We thus propose a synergistic pathophysiological model in COMISA. In this model, OSA-related symptoms, particularly intermittent hypoxia and recurrent sleep fragmentation, are exacerbated in the presence of insomnia, amplifying downstream effects on the autonomic nervous system, oxidative stress, and cardiovascular remodeling[41]. Likewise, the multiplicative risk of chronic muscular pain and headache in COMISA may be also caused by the synergistic effect of insomnia and OSA on sleep fragmentation and intermittent hypoxia. Current evidence suggests that the existence of a bidirectional relationship between sleep disturbances and chronic pain[53]. The findings from the Sleep Heart Health Study indicates that a reduction in PSG derived TST is associated with increased moderate to severe pain[53, 54]. The coexistence of insomnia and OSA could jointly affect negatively objective sleep, notably its continuity, thereby further causing chronic pain.

Interestingly, we found a synergistic effect of pre-sleep cognitive-emotional and somatic hyperarousal between insomnia and OSA. While OSA alone decreases the risk of pre-sleep symptoms such as falling asleep anxious, depressed etc, in combination and for AHI above 15, combined it results in an exacerbation of pre-sleep complaints. Consistent with our findings, a previous study showed that patients with COMISA report significantly higher pre-sleep anxiety, depression, and arousal than those with OSA alone[43], indicating that the coexistence of insomnia and OSA gives rise to a synergistic phenotype of psychological hyperarousal. It is suggested that individuals with subjective EDS have an increased self-awareness with their own sleep health issues, which may be due to increased psychological stress[50]. Psychological hyperarousal is widely recognized as a core mechanism in the pathophysiology of insomnia[55]. The additional risk of subjective EDS in COMISA may be caused by the synergistic effect of insomnia and OSA on psychological hyperarousal.

In this study, we found no evidence that gender modifies the joint association of insomnia and OSA with cardiovascular and neuropsychological comorbidities or pre-sleep symptoms. However, we observed that in men, the adverse effects of insomnia and OSA were amplified when the other condition was present, indicating a male-specific vulnerability in associating respiratory disturbance and REM sleep decreasing. Several mechanisms may underlie a gender difference. Men have greater upper-airway collapsibility and ventilatory instability than women[14, 56–58]. At the same time, insomnia-associated hyperarousal may further exacerbate OSA-related sleep fragmentation in men, producing a more pronounced bidirectional reinforcement between cortical arousal and respiratory events[1].

### Strengths and Limitations

This study has important strengths. It draws on a large, multi-center clinical data, providing robust statistical power to detect interactions that were often overlooked in prior COMISA research. The simultaneous evaluation of multiplicative and additive interactions across a comprehensive range of comorbidities, sleep symptoms and PSG parameters offers granular insight into the heterogeneity and clinical complexity of COMISA. Further, to control for false positive findings, we conducted FDR corrections, making the interpretation of the results more rigorous. Several limitations should however be acknowledged. First, our interaction analyses derive from observational cross-sectional data, limiting causal inference. Longitudinal or interventional studies will be needed to clarify whether the observed interactions represent causal pathways or bidirectional reinforcement. Second, comorbidities in this study were assessed through self-reported questionnaires rather than standardized clinical diagnoses, which may introduce reporting bias. Future work incorporating objective clinical evaluations and validated diagnostic instruments is needed to strengthen the reliability of these findings. Third, the baseline PSGs in this analysis were collected over a very long time period (2004 to 2019). To address it, we conducted a sensitivity analysis adding 5 years blocks of referral as a covariate and found it to be non-confounding. Finally, the sample size is large, and some of the interactions noted, although significant, are of modest magnitude.

## Conclusion

Our findings highlight COMISA as a distinct and clinically high-risk phenotype characterized by the amplification of both biological disturbances (sleep fragmentation and intermittent hypoxia) and psychological hyperarousal.

## Data Availability

Summary data is available upon request

## Acknowledgments

Author contributions: E.M. serves as the guarantor of the paper, taking responsibility for the integrity of the work, from inception to publication of the article. The study was designed by E.M., Y.D., and U.H.; data were collected by U.H. and U.G.; data were analyzed by Y.D., and results interpreted by all authors. The manuscript and figures were drafted by Y.D. and critically revised by all authors.

## Data availability statement

Summary data is available upon request.

---

**Figure S1.**
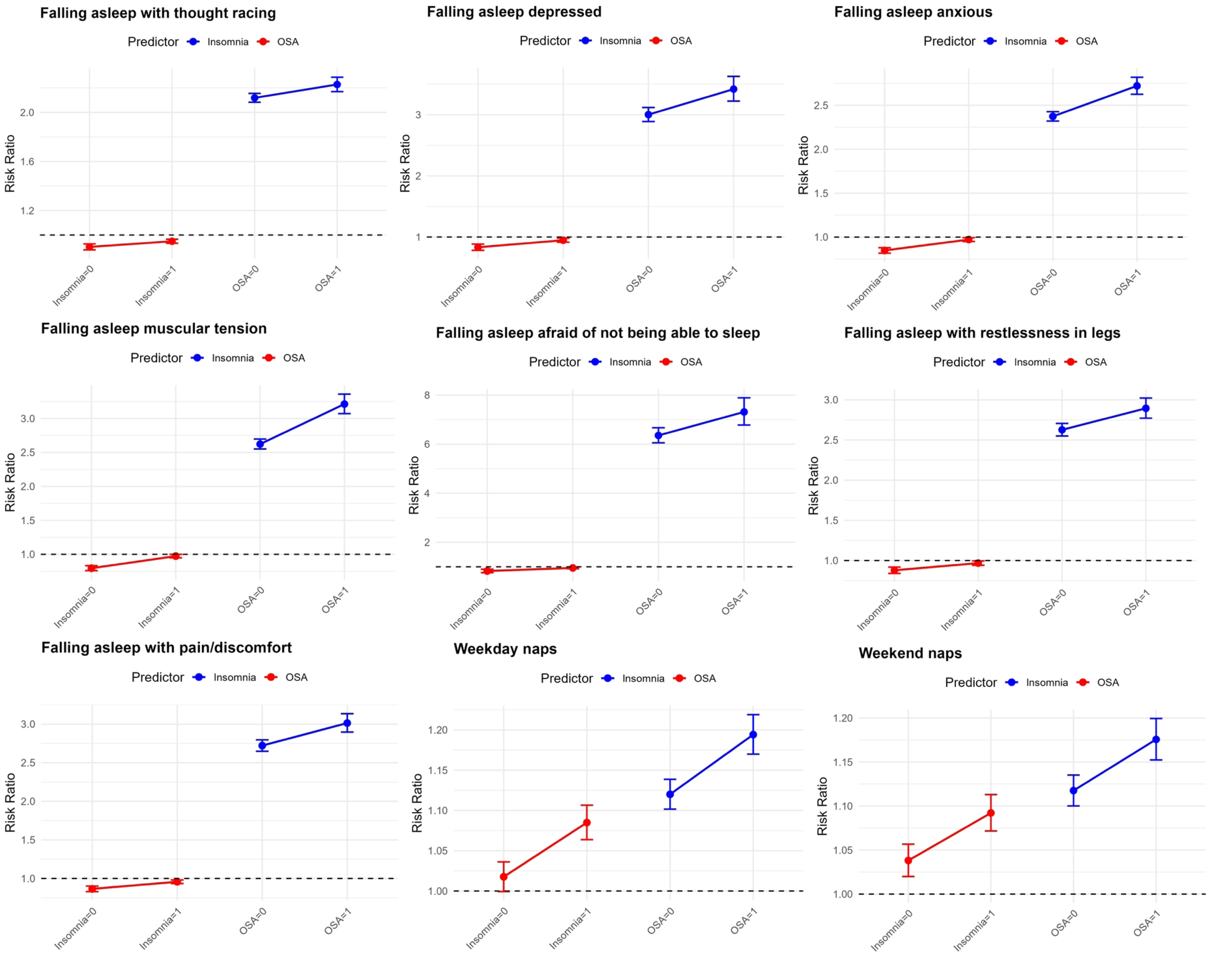
Simple slopes analyses of the interactions between insomnia and OSA on comorbidities. Abbreviations: EDS, excessive daytime sleepiness; OSA, obstructive sleep apnea.

**Figure S2.**
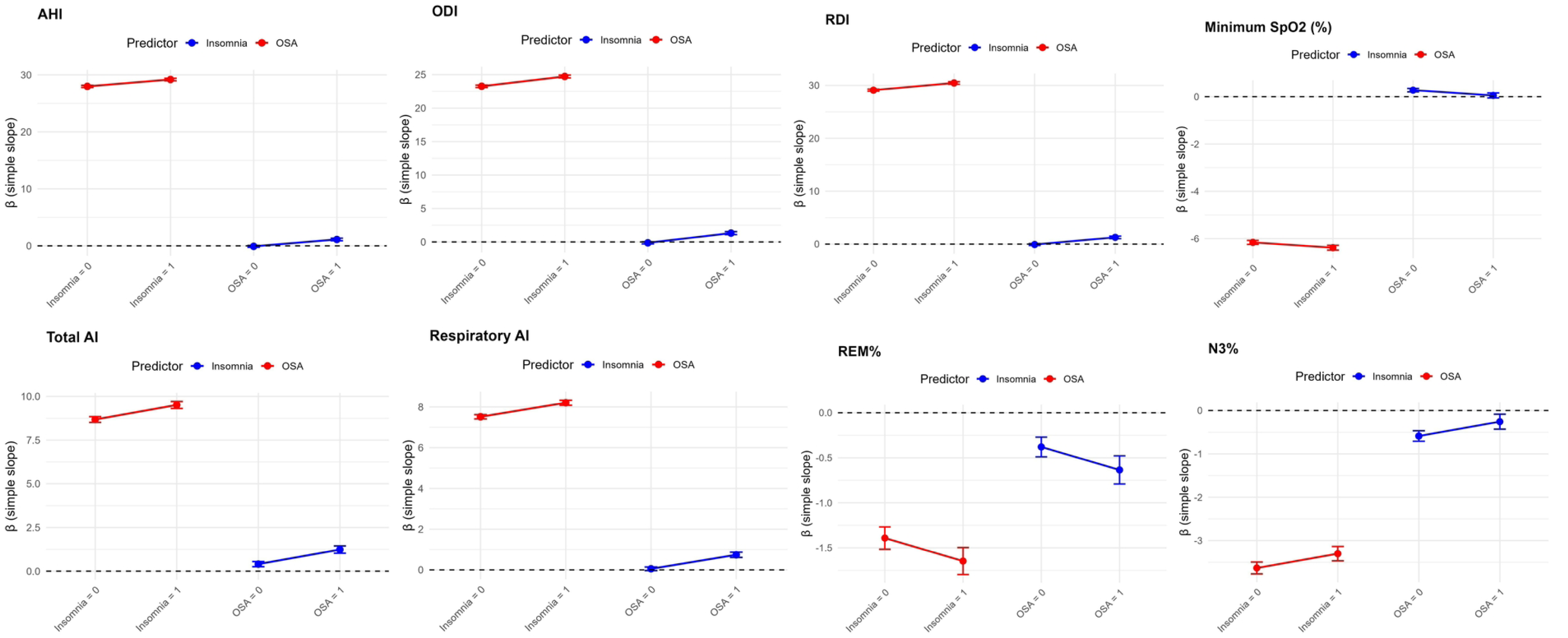
Simple slopes analyses of the interactions between insomnia and OSA on sleep symptoms. Abbreviations: OSA, obstructive sleep apnea.

**Figure S3.**
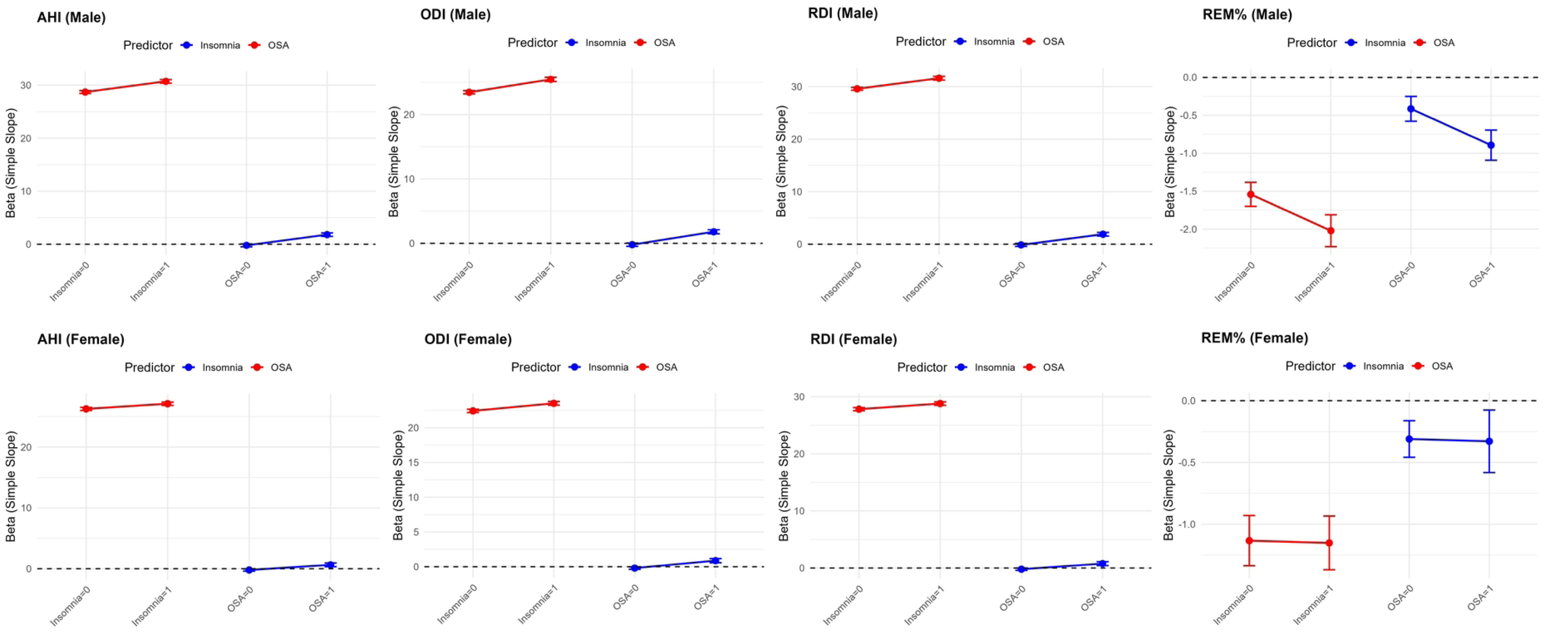
Simple slopes analyses of the interactions between insomnia and OSA on PSG parameters. Abbreviations: AHI, Apnea Hypopnea Index; AI, arousal index; N3%, percentage of non-rapid eye movement sleep stage 3; ODI, oxygen desaturation index; OSA, obstructive sleep apnea; RDI, respiratory disturbance index; REM%, percentage of rapid eye movement sleep stage; SpO_2_, oxygen saturation.

**Figure S4.** Forest plot of the three-way interactions between insomnia, OSA and gender on comorbidities, sleep symptoms and PSG parameters. Note: A. Forest plot of the interactions between insomnia, OSA and male on comorbidities and sleep symptoms; B. Forest plot of the interactions between insomnia, OSA and male on PSG parameters. Abbreviations: AHI, Apnea Hypopnea Index; AI, arousal index; EDS, excessive daytime sleepiness; N3%, percentage of non-rapid eye movement sleep stage 3; ODI, oxygen desaturation index; OSA, obstructive sleep apnea; RDI, respiratory disturbance index; REM%, percentage of rapid eye movement sleep stage; SpO_2_, oxygen saturation.

**Figure S5.** Gender-stratified simple slopes analyses of the interactions between insomnia and OSA on PSG parameters. Abbreviations: AHI, Apnea Hypopnea Index; N3%, percentage of non-rapid eye movement sleep stage 3; ODI, oxygen desaturation index; OSA, obstructive sleep apnea; RDI, respiratory disturbance index; REM%, percentage of rapid eye movement sleep stage.

**Table S1.** Main effects and multiplicative interactions between insomnia and OSA on comorbidities and sleep symptoms.

| Outcome | Insomnia RR (95% CI) | Adjusted P value | OSA RR (95% CI) | Adjusted P value | Interaction RR (95% CI) | Adjusted P value |
| --- | --- | --- | --- | --- | --- | --- |
| **Comorbidities** |  |  |  |  |  |  |
| High blood pressure | 1.069 (1.059, 1.079) | **2.7e<sup>-43</sup>** | 1.095 (1.085, 1.106) | **1.6e<sup>-76</sup>** | 0.995 (0.977, 1.014) | 0.808 |
| Diabetes | 1.130 (1.108, 1.151) | **6.4e<sup>-36</sup>** | 1.091 (1.070, 1.113) | **4.9e<sup>-18</sup>** | 0.975 (0.940, 1.012) | 0.337 |
| Heart attack | 1.287 (1.240, 1.335) | **8.0e<sup>-41</sup>** | 1.081 (1.041, 1.122) | **6.9e<sup>-5</sup>** | 0.995 (0.925, 1.069) | 0.883 |
| Stroke | 1.203 (1.156, 1.251) | **4.7e<sup>-20</sup>** | 0.988 (0.948, 1.030) | 0.574 | 1.017 (0.939, 1.101) | 0.808 |
| Heart enlargement | 1.345 (1.283, 1.410) | **1.1e<sup>-34</sup>** | 1.043 (0.993, 1.095) | 0.101 | 1.110 (1.011, 1.218) | 0.062 |
| Irregular heartbeat | 1.320 (1.294, 1.347) | **2.0e<sup>-160</sup>** | 0.943 (0.923, 0.964) | **1.9e<sup>-7</sup>** | 0.994 (0.953, 1.036) | 0.832 |
| Depression | 1.527 (1.500, 1.555) | **< 10e<sup>-300</sup>** | 0.939 (0.921, 0.958) | **6.3e<sup>-10</sup>** | 1.068 (1.029, 1.110) | **0.003** |
| Memory loss | 1.613 (1.578, 1.649) | **< 10e<sup>-300</sup>** | 0.895 (0.873, 0.917) | **7.0e<sup>-19</sup>** | 1.026 (0.980, 1.075) | 0.394 |
| Heartburn | 1.337 (1.319, 1.356) | **< 10e<sup>-300</sup>** | 1.017 (1.002, 1.033) | **0.031** | 1.032 (1.003, 1.061) | 0.062 |
| Chronic muscular pain | 1.755 (1.722, 1.789) | **< 10e<sup>-300</sup>** | 0.928 (0.909, 0.947) | **9.6e<sup>-13</sup>** | 1.058 (1.017, 1.101) | **0.017** |
| Headache | 1.546 (1.523, 1.570) | **< 10e<sup>-300</sup>** | 0.878 (0.862, 0.894) | **4.0e<sup>-43</sup>** | 1.121 (1.082, 1.160) | **1.1e<sup>-9</sup>** |
| Nocturia | 1.794 (1.767, 1.820) | **< 10e<sup>-300</sup>** | 1.064 (1.048, 1.080) | **1.7e<sup>-15</sup>** | 0.981 (0.953, 1.010) | 0.337 |
| Subjective EDS | 1.376 (1.360, 1.392) | $< 10e^{-300}$ | 1.072 (1.059, 1.086) | $1.6e^{-26}$ | 1.089 (1.063, 1.115) | $1.6e^{-11}$ |
| **Sleep Symptoms** |  |  |  |  |  |  |
| Falling asleep with<br>thought racing | 2151 (2.120, 2.183) | $< 10e^{-300}$ | 0.931 (0.917, 0.945) | $2.2e^{-19}$ | 1.051 (1.019, 1.084) | **0.002** |
| Falling asleep<br>depressed | 3.120 (3.020, 3.224) | $< 10e^{-300}$ | 0.913 (0.885, 0.942) | $2.0e^{-8}$ | 1.139 (1.063, 1.220) | $2.9e^{-4}$ |
| Falling asleep<br>anxious | 2.470 (2.423, 2.132) | $< 10e^{-300}$ | 0.928 (0.910, 0.946) | $1.8e^{-13}$ | 1.146 (1.100, 1.195) | $3.9e^{-10}$ |
| Falling asleep<br>muscular tension | 2.781 (2.714, 2.849) | $< 10e^{-300}$ | 0.915 (0.894, 0.937) | $4.5e^{-13}$ | 1.225 (1.163, 1.289) | $1.1e^{-13}$ |
| Falling asleep afraid<br>of not being able to<br>sleep | 6.623 (6.354, 6.904) | $< 10e^{-300}$ | 0.932 (0.904, 0.961) | $8.2e^{-6}$ | 1.151 (1.054, 1.257) | **0.002** |
| Falling asleep with<br>restlessness in legs | 2.709 (2.642, 2.778) | $< 10e^{-300}$ | 0.937 (0.914, 0.960) | $2.2e^{-7}$ | 1.102 (1.047, 1.160) | $2.9e^{-4}$ |
| Falling asleep with<br>pain/discomfort | 2.812 (2.749, 2.877) | $< 10e^{-300}$ | 0.927 (0.907, 0.947) | $1.0e^{-11}$ | 1.107 (1.057, 1.160) | $4.5e^{-5}$ |
| Weekday naps | 1.146 (1.131, 1.161) | **1.2e<sup>-92</sup>** | 1.046 (1.032, 1.061) | **1.4e<sup>-10</sup>** | 1.066 (1.039, 1.094) | **3.9e<sup>-6</sup>** |
| Weekend naps | 1.138 (1.124, 1.152) | **1.9e<sup>-90</sup>** | 1.061 (1.047, 1.076) | **1.0e<sup>-17</sup>** | 1.052 (1.026, 1.079) | **1.3e<sup>-4</sup>** |

Note: Modified Poisson regression models were adjusted for age, gender, BMI, race, tobacco use, alcohol consumption, caffeinated beverage use, and daily use of sleep aids. Multiplicative interaction was assessed by including the interaction term Insomnia×OSA in models. Multiple comparisons were addressed by controlling the false discovery rate. Outcomes were grouped into comorbidities, sleep symptoms and PSG parameters families, and p values within each family were adjusted using the Benjamini-Hochberg method.
Abbreviations: CI, confidence interval; EDS, excessive daytime sleepiness; OSA, obstructive sleep apnea; RR, risk ratio.

**Table S2.**
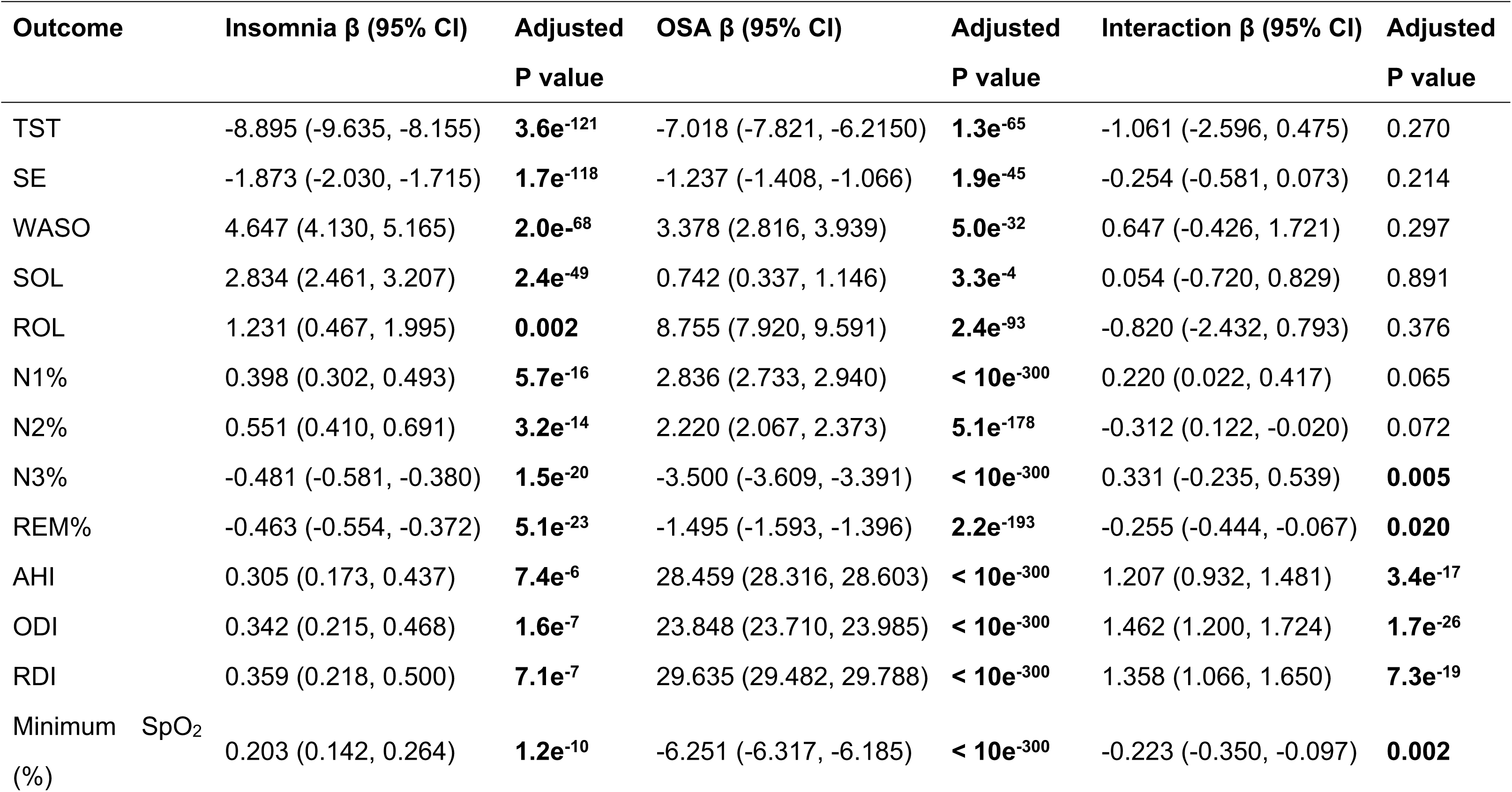
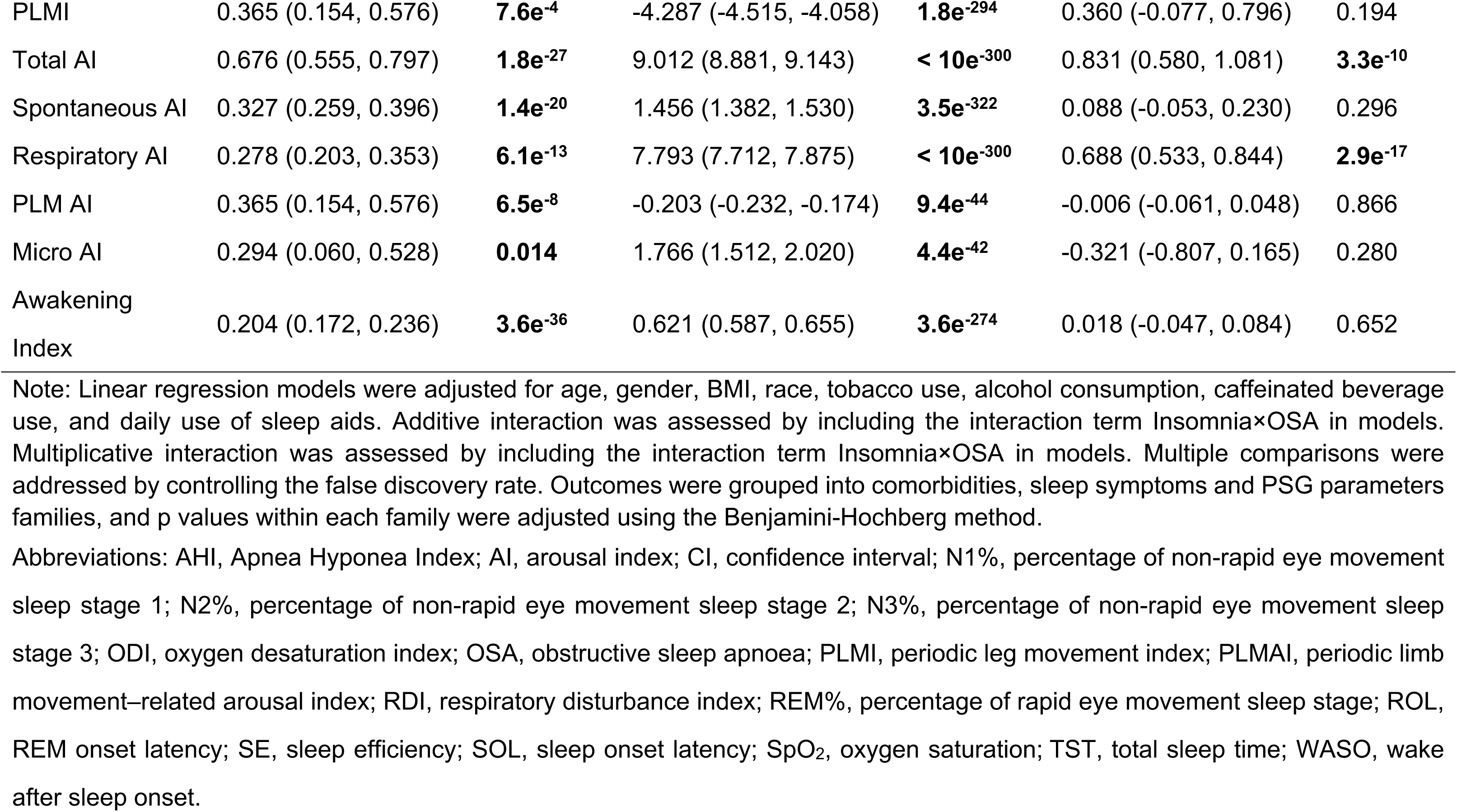
Main effects and additive interactions between insomnia and OSA on PSG parameters.

**Table S3.**
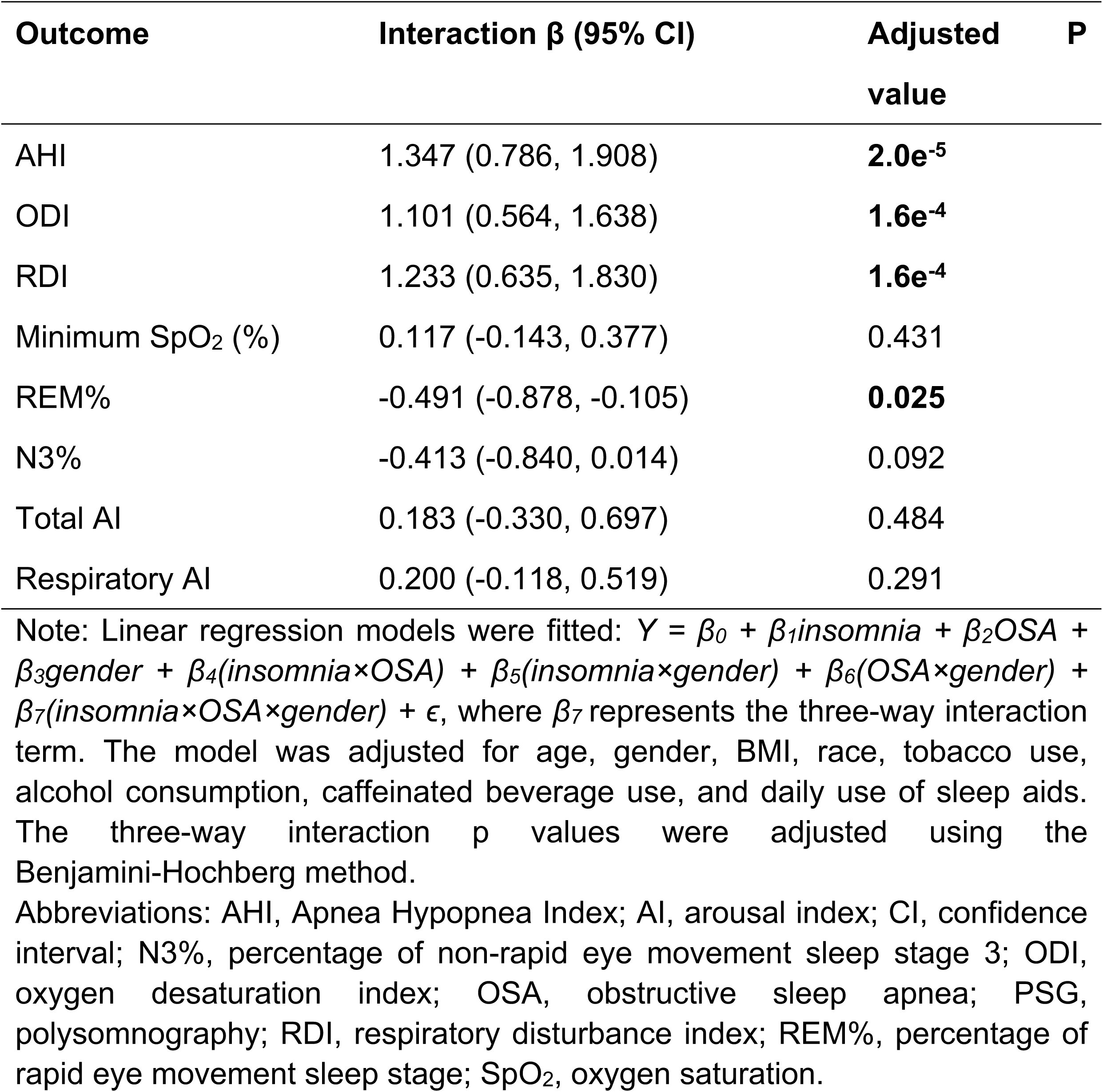
Additive interactions between insomnia, OSA and gender (male) on PSG parameters.

